# Parkinson’s Families Project: a UK-wide study of early onset and familial Parkinson’s disease

**DOI:** 10.1101/2023.12.05.23299397

**Authors:** Clodagh Towns, Zih-Hua Fang, Manuela M. X. Tan, Simona Jasaityte, Theresa M. Schmaderer, Eleanor J. Stafford, Miriam Pollard, Russel Tilney, Megan Hodgson, Lesley Wu, Robyn Labrum, Jason Hehir, James Polke, Lara M. Lange, Anthony H. V. Schapira, Kailash P. Bhatia, Parkinson’s Families Project (PFP) Study Group, Global Parkinson’s Genetics Program (GP2), Andrew B. Singleton, Cornelis Blauwendraat, Christine Klein, Henry Houlden, Nicholas W. Wood, Paul R. Jarman, Huw R. Morris, Raquel Real

## Abstract

The Parkinson’s Families Project is a UK-wide study aimed at identifying genetic variation associated with familial and early-onset Parkinson’s disease (PD). We recruited individuals with a clinical diagnosis of PD and age at motor symptom onset ≤ 45 years and/or a family history of PD in up to third-degree relatives. Where possible, we also recruited affected and unaffected relatives. We analysed DNA samples with a combination of single nucleotide polymorphism (SNP) array genotyping, multiplex ligation-dependent probe amplification (MLPA), and whole-genome sequencing (WGS). We investigated the association between identified pathogenic mutations and demographic and clinical factors such as age at motor symptom onset, family history, motor symptoms (MDS-UPDRS) and cognitive performance (MoCA). We performed baseline genetic analysis in 718 families, of which 205 had sporadic early-onset PD (sEOPD), 113 had familial early-onset PD (fEOPD), and 400 had late-onset familial PD (fLOPD). 69 (9.6%) of these families carried pathogenic variants in known monogenic PD-related genes. The rate of a molecular diagnosis increased to 28.1% in PD with motor onset ≤ 35 years. We identified pathogenic variants in *LRRK2* in 4.2% of families, and biallelic pathogenic variants in *PRKN* in 3.6% of families. We also identified two families with *SNCA* duplications and three families with a pathogenic repeat expansion in *ATXN2*, as well as single families with pathogenic variants in *VCP*, *PINK1*, *PNPLA6*, *PLA2G6*, *SPG7*, *GCH1*, and *RAB32*. An additional 73 (10.2%) families were carriers of at least one pathogenic or risk *GBA1* variant. Most early-onset and familial PD cases do not have a known genetic cause, indicating that there are likely to be further monogenic causes for PD.

## INTRODUCTION

Parkinson’s disease (PD) is the second most common neurodegenerative condition after Alzheimer’s Disease (AD) and its prevalence is rapidly increasing^1^. PD becomes more common with advancing age, and both common and rare genetic variants can increase the risk of PD. Additionally, rare variants in approximately 20 genes have been reported to cause monogenic PD, although some of these genes have not been widely replicated, and some cause syndromes that are clinically and/or pathologically distinct from sporadic late-onset PD (sLOPD)^2,3^. First-degree relatives of PD patients have been estimated to have an approximately 2-fold increased risk of developing the condition compared to unrelated individuals^4–6^. A family history of PD and an early age at onset (AAO) are associated with an increased likelihood of carrying a pathogenic variant^7,8^. In unselected PD populations, rare causal variants account for around 1-2% of patients, whereas rare causal variants are found in around 5% of patients with familial PD and 20-40% of patients with an age of onset ≤30^9^. Pathogenic variants in *LRRK2*, *SNCA* and *VPS35* have been consistently identified in autosomal dominant PD, and biallelic pathogenic variants in *PRKN, PINK1*, *DJ-1*, and *ATP13A2* in autosomal recessive PD. Recently, a single pathogenic variant in *RAB32* has been identified in autosomal dominant families^10,11^. Rare variants in the Gaucher disease-causing *GBA1* gene are an important genetic risk factor for PD, with approximately 5-10 % of Northern European PD patients carrying single *GBA1* variants^12^. For the vast majority of early-onset and familial PD cases, a known genetic cause has not been identified, suggesting either that there are additional monogenic forms to discover and/or that some PD families have more complex inheritance^13,14^.

Global efforts are underway to collect clinical and genetic data of diagnosed PD cases to elucidate the multifactorial pathogenesis of this complex disease^15–20^. However, a major obstacle to identifying and validating candidate monogenic variants is the availability of DNA samples from affected and unaffected family members. Classic linkage analysis and whole-exome/genome sequencing strategies have been used to show a causal relationship between genetic variation and monogenic PD, both of which require access to DNA samples from multiple family members across several generations^21^.

The Parkinson’s Families Project (PFP) is an ongoing UK-wide study aiming to identify new monogenic forms of PD by recruiting PD patients who are more likely to have a strong genetic contribution to the development of the condition, as well as their affected and unaffected relatives. UK-based studies of PD have previously shown that early-onset PD (EOPD) with age at symptom onset <45 years, as well as PD families with three or more affected members are particularly likely to carry a pathogenic mutation^8^. Here, we have built on this approach by recruiting early-onset and/or familial PD cases together with their genetically related family members to enable further genetic investigation of PD. The aims of the PFP study are: i) to build a cohort of families in which new monogenic variants may be discovered, and candidate pathogenic variants may be replicated through segregation studies; ii) to define the frequency and clinical features of pathogenic variants in known PD genes in a large-scale multicentre study; iii) to define a cohort of patients eligible for precision drug trials. PFP started recruitment in 2015 and will continue to do so until January 2030, with a target recruitment of over 1,500 families, comprising over 3,000 participants. Here, we describe the study protocol and the preliminary findings from our genetic screening of the first 718 families.

## RESULTS

### Cohort description

We recruited 1,035 participants from 840 families to the PFP study. Of these, we evaluated 959 individuals from 785 families using at least one of the genetic testing techniques described below. We then excluded 67 index cases from further analysis due to either a diagnosis of secondary parkinsonism (*n* = 3), atypical parkinsonism (*n* = 6), non-parkinsonism disorder (*n* = 2), failure to meet inclusion criteria (*n* = 30), missing clinical data (*n* = 14), consent withdrawal (*n* = 1), duplicated samples (*n* = 6), or failed genetic testing (*n* = 5) (Supplementary Figure 1). Relatives of excluded index cases were also excluded (*n* = 16 relatives). In total, data were available from 871 eligible participants from 718 families.

Baseline demographics and PD family history for the 718 index cases included are shown in Table 1. 28.6% (205/718) of index cases have sporadic early-onset PD (sEOPD), 15.7% (113/718) have familial early-onset PD (fEOPD), and 55.7% (400/718) have familial late-onset PD (fLOPD). Using genetic principal component analysis (PCA) to define ancestry, 92.8% of all index cases were of European ancestry. In most families only the index case was recruited, but in 16% (*n* = 117) at least one additional relative was also recruited. Kinship analysis identified four families with cryptic relatedness. In all of these cases, individuals from the same extended family were independently recruited at different study sites. Across all families, we recruited 37 affected and 116 unaffected relatives for segregation studies. Of these multiplex families, 72% consisted of the index case and one single relative, while 20% had two relatives recruited, and 8% had three or more relatives recruited. For 7.9% of early-onset index cases, at least one parent was recruited. In all but one family, we recruited only a single additional affected relative.

**Table 1.**
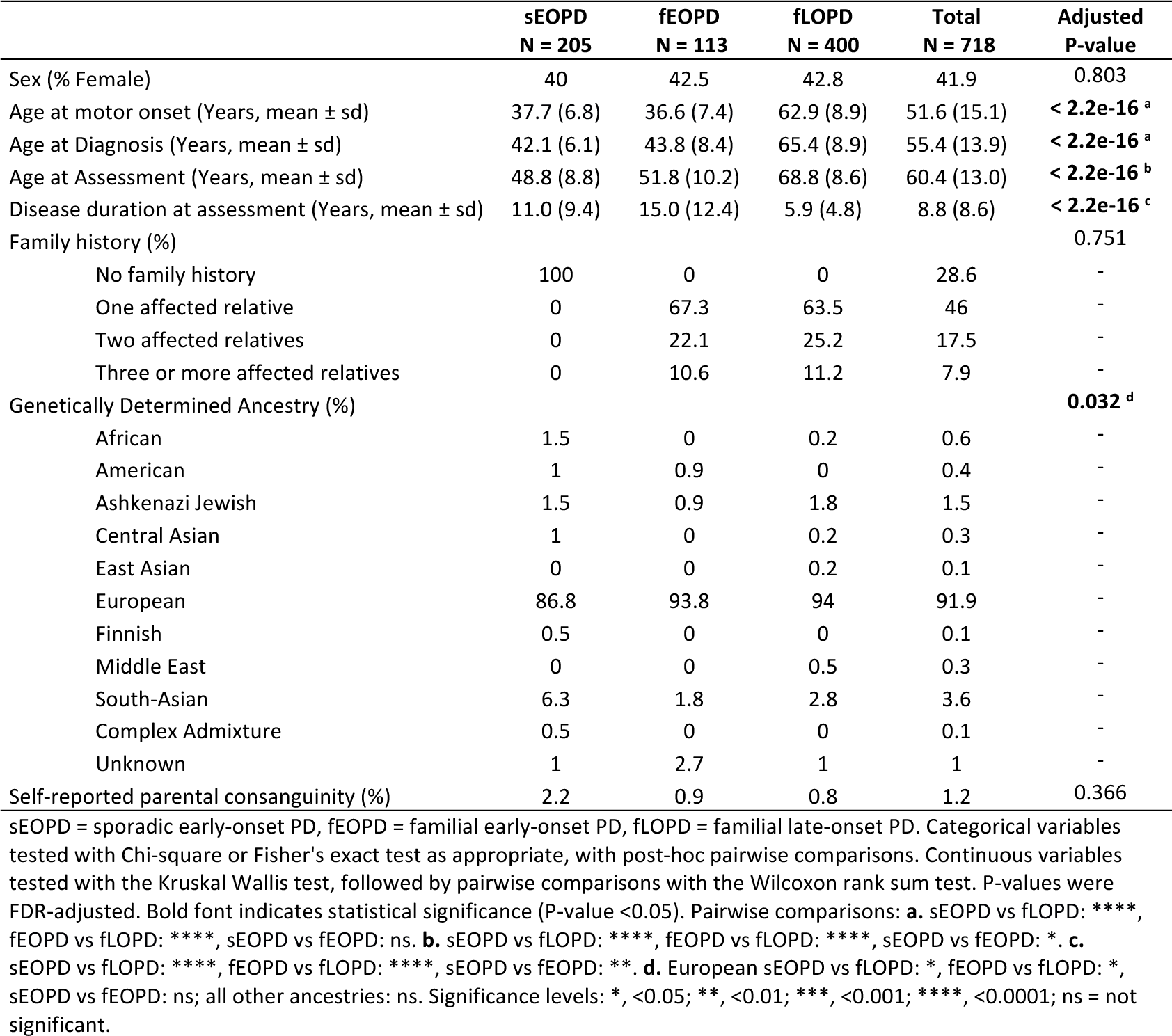
Demographic characteristics of index cases by group.

### Identification of PD-causing variants

Following completion of genetic analysis by a combination of Illumina’s NeuroChip genotyping array (NCA), multiplex ligation-dependent probe amplification (MLPA), whole-genome sequencing (WGS) and/or next-generation targeted sequencing (NGS), we identified known PD-causing variants in 69 families (9.6%, 69/718; Supplementary Table 1). NCA contains probes for hundreds of PD relevant rare variants. We tested the performance of these probes against whole-genome or targeted sequencing (Supplementary Methods and Supplementary Table 2) and found that NCA-derived genotypes showed 95.2% concordance with sequenced-derived genotypes, indicating a high level of accuracy for most probes. Poorly performing probes were excluded from subsequent analyses.

Rare pathogenic variants in autosomal dominant genes explained PD occurrence in 38 families (5.3%; Table 2 and Supplementary Table 3). Mutations in *LRRK2* were the most commonly identified genetic cause, accounting for PD in 30 families (4.2%). The *LRRK2* G2019S variant was identified in all but two of these families. The majority (*n* = 23; 76.7%) of the *LRRK2* mutation-positive families had fLOPD. Interestingly, five *LRRK2* G2019S carriers had sEOPD, reflecting incomplete penetrance and the likely presence of disease modifiers. Other pathogenic dominant variants identified include two cases of heterozygous *SNCA* gene duplication and three cases with expanded trinucleotide repeats in *ATXN2*. *SNCA* copy number variants (CNVs) are typically associated with fEOPD^22^, but both these cases presented as sEOPD. We have also identified pathogenic missense variants in *VCP*, *GCH1* and *RAB32*. The *RAB32* p.Ser71Arg here identified has recently been reported in several autosomal dominant PD families, and has been shown to activate LRRK2 kinase *in vitro*^10,11^.

**Table 2.**
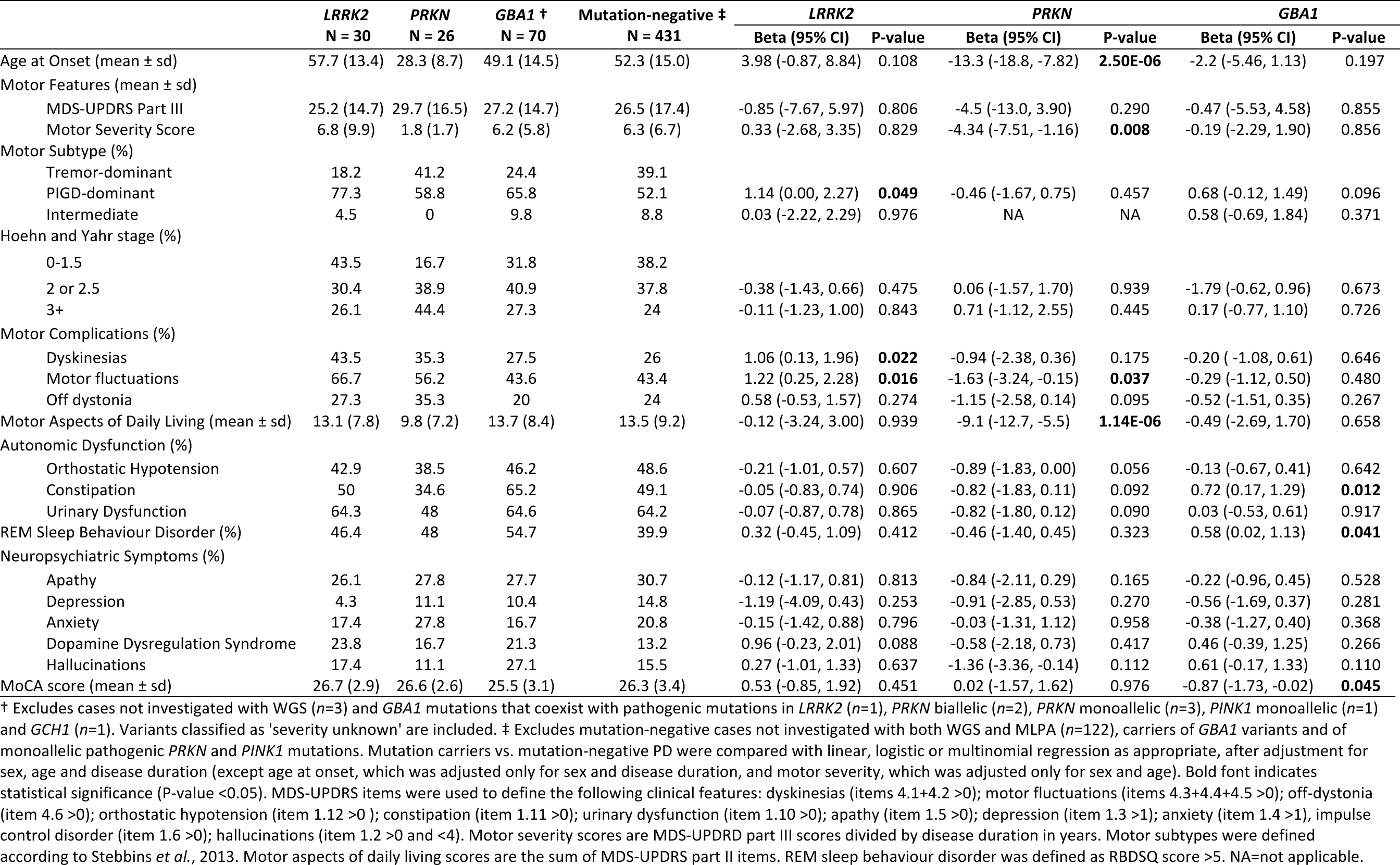
Clinical features of mutation carriers vs. mutation-negative index cases.

Pathogenic biallelic autosomal recessive variants were identified in 31 families (4.3%; Table 2 and Supplementary Table 3). Compound heterozygous or homozygous pathogenic variants in *PRKN* were the second most common cause of monogenic PD, accounting for PD in 26 families (3.6%). All biallelic *PRKN* index cases presented with early-onset PD, and 15 (57.7%) cases did not have a family history of PD. Consanguinity was reported in 4.3% of biallelic *PRKN* mutation carriers compared to 1.2% of early-onset PD cases without mutations (P = 0.298, Fisher’s Exact test). *PINK1* homozygous pathogenic variants were identified in two families. The remaining biallelic recessive cases carried homozygous variants in *PNPLA6*, and compound heterozygous variants in *PLA2G6* and *SPG7*.

Supplementary Table 1 lists genetic findings of all participants with known PD-causing variants and their relatives. We further identified 23 index cases with a single heterozygous pathogenic variant in either *PRKN* or *PINK1*, of whom 14 were fully investigated with WGS and MLPA (Supplementary Table 4). A list of all the unique variants identified (*n* = 72, including *GBA1* risk variants) is provided in Supplementary Table 5.

### Demographic characteristics of pathogenic variant carriers

As expected, pathogenic variants in PD-related genes were more common in participants with an early AAO, defined by symptom onset before age 45 years (Supplementary Table 6). We identified a monogenic cause in 12.9% (41/318) of patients with EOPD (≤ 45 years) compared to 7% (28/400) of patients with LOPD (ꭓ^2^ = 7.1, df = 1, 95% confidence interval [CI] = 0.01-0.10, P = 0.008, Chi-squared test). Moreover, when looking into juvenile and young onset PD (≤ 35 years), a monogenic cause was present in 28.1% (27/96) of patients with symptom onset ≤ 35, compared to 6.7% (42/622) of patients with AAO > 35 (ꭓ2 = 43.7, df = 1, 95%CI = 0.12-0.31, P = 3.76e-11, Chi-squared test). In particular, 26% (25/96) of patients with symptom onset ≤ 35 carried homozygous or compound heterozygous mutations in recessive genes, compared to only 0.96% in patients with onset > 35 (6/622; P = 2.2e-16, Fisher’s exact test). Among patients with a family history of PD, dominant mutations were more frequent than biallelic recessive mutations (6.0% vs 2.5%; ꭓ2 = 6.9, df = 1, 95%CI = 0.01-0.06, P = 0.009, Chi-squared test). Furthermore, each additional affected family member increased the odds of having a dominant mutation by a factor of 1.6, after adjusting the logistic regression for sex and age at symptom onset (95%CI = 1.21-2.02, P = 5.34e-04). The majority of pathogenic mutation carriers were of European ancestry, except for one participant of South East Asian ancestry with homozygous pathogenic mutations in *PINK1* (Y258*), and four participants of Ashkenazi Jewish ancestry (three heterozygous *LRRK2* G2019S carriers and one homozygous *PNPLA6* P1297S carrier).

### Clinical features of *LRRK2* mutation carriers

Among *LRRK2* mutation carriers, 83.3% (25/30) had a positive family history of PD, and the majority experienced symptom onset > 45 years (76.7%, 23/30). Demographic characteristics of *LRRK2* mutation carriers are described in Supplementary Table 7. Clinical features of PD-*LRRK2* mutation carriers compared to mutation-negative index cases (i.e., no identified dominant or biallelic/monoallelic recessive variants in PD-related genes or *GBA1*) are presented in Table 2. Age at onset was similar in PD-*LRRK2* and mutation-negative PD (57.7 ± 13.4 vs 52.3 ± 15.0 years; r= 0.09, P = 0.063, Mann-Whitney U test). While the majority of *LRRK2* mutation carriers were European, 10% were of Ashkenazi Jewish ancestry compared to 0.91% of mutation-negative PD (P = 0.006, Fisher’s exact test). We compared the PD motor subtype in PD-*LRRK2* and mutation-negative PD using multinomial logistic regression, adjusted for sex, age, and disease duration. PD-*LRRK2* cases had an increased odds ratio (OR) of having a postural instability and gait difficulty (PIGD)-dominant compared to a tremor-dominant motor subtype (OR = 3.1, 95%CI = 1.00-9.71, P = 0.049). There was no difference in motor severity, as measured by MDS-UPDRS part III, between PD-*LRRK2* and mutation-negative PD (25.2 ± 14.7 vs 26.5 vs 17.4, respectively; r = 0.01, P = 0.821, Mann-Whitney U test). Regarding motor complications, motor fluctuations were more common in PD-*LRRK2* (Chi-squared test: ꭓ^2^ = 4.2, df = 1, 95%CI = 0.02-0.44, P = 0.039) and there was also a tendency towards a higher rate of dyskinesia in PD-LRRK2 (Chi-squared test: ꭓ^2^ = 3.2, df = 1, 95%CI = -0.03-0.38, P = 0.071). We then adjusted for sex, age, and disease duration in a logistic regression model, which confirmed the association between *LRRK2* mutations and dyskinesia and motor fluctuations (dyskinesia: OR = 2.9, 95%CI = 1.14-7.12, P = 0.022; motor fluctuations: OR = 3.4, 95%CI = 1.29-9.75, P = 0.016). No other comparisons of clinical features between PD-*LRRK2* and mutation-negative PD cases reached significance.

### Clinical features of biallelic *PRKN* mutation carriers

The demographic and clinical features of biallelic *PRKN* mutation carriers are summarised in Supplementary Table 7 and Table 2, respectively. 42.3% (11/26) of biallelic *PRKN* mutation carriers had a positive family history of PD. The majority had symptom onset ≤ 35 years (80.8%, 21/26), while 23.1% (6/26) had juvenile PD (i.e., symptom onset ≤ 21). Accordingly, biallelic *PRKN* mutation carriers had a significantly earlier age of symptom onset compared to mutation-negative PD (28.3 ± 8.7 vs 52.3 ± 15.0 years; r = 0.34, P = 5.49e-13, Mann-Whitney U test). Disease duration was also significantly longer at study assessment (21.7 ± 14.0 vs 8.37 ± 8.31; r = 0.25, P = 8.34e-08, Mann-Whitney U test). All biallelic *PRKN* mutation carriers were of European ancestry. There were no differences in motor scores or motor subtypes between groups. However, given that biallelic *PRKN* mutation carriers had significantly longer disease duration, we adjusted motor severity to disease duration by dividing MDS-UPDRS part III scores at assessment by disease duration. Biallelic *PRKN* mutation carriers had significantly lower adjusted motor severity scores compared to PD without a monogenic cause (1.8 ± 1.7 vs 6.3 ± 6.7; r = 0.26, P = 1.76e-05, Mann-Whitney U test), indicating a slower rate of motor symptom progression. Concordantly, individuals with biallelic *PRKN* mutations performed better in motor aspects of activities of daily living, as measured by MDS-UPDRS part II, after adjusting for confounding variables including disease duration (linear regression: beta = -9.1, standard error (sd) = 1.8, P = 1.14e-06). Biallelic *PRKN* carriers had increased rate of motor fluctuations at baseline (56.2% vs 30.9%; ꭓ^2^ = 4.5, df = 1, 95%CI = 0.00-0.50, P = 0.0339, Chi-squared test). However, biallelic *PRKN* mutations were associated with a reduced likelihood of experiencing motor fluctuations compared to mutation-negative PD after adjusting for disease duration (OR = 0.20, 95%CI = 0.04-0.91, P = 0.0369). No other clinical features differentiated biallelic *PRKN* mutation carriers from mutation-negative PD cases. In addition to biallelic *PRKN* mutation carriers, there were 12 index cases fully investigated with WGS and MLPA for whom only a single pathogenic mutation could be found. Interestingly, monoallelic *PRKN* pathogenic variant carriers were more similar to mutation-negative PD than to biallelic *PRKN* pathogenic variant carriers (Supplementary Table 8).

### Demographic and clinical features of pathogenic and risk *GBA1* variant carriers

We screened *GBA1* for rare pathogenic Gaucher disease (GD)-causing variants and common PD risk variants. We identified 73 carriers of *GBA1* variants and an additional eight index cases with concomitant pathogenic mutations in *LRRK2*, *PRKN*, *PINK1* and *GCH1* (Supplementary Table 9). 3.7% (3/81) of *GBA1* carriers were of Ashkenazi Jewish ancestry. After excluding *GBA1* carriers with coexistent pathogenic mutations in other PD-related genes (*n*=8) and those who did not complete WGS (*n*=3), 70 individuals were available for subsequent analysis (Table 2). A family history of PD was present in 71.4% (50/70) of *GBA1* mutation carriers, and in 93.9% of these families affected individuals were present in at least two generations. 50% (35/70) of *GBA1* mutation carriers had motor symptom onset ≤ 45 years, in line with previous studies suggesting earlier symptom onset in *GBA1* mutation carriers^23,24^. Compared to mutation-negative PD, *GBA1* mutation carriers had decreased MoCA scores after adjusting for age at assessment and disease duration (beta = -0.87, sd = 0.43, P = 0.045). In addition, the odds of REM sleep behaviour disorder, which is often a precursor of cognitive decline and dementia in PD^25–27^, were significantly increased in *GBA1* carriers (OR = 1.79, 95%CI = 1.02-3.11, P = 0.041). We also found an association between constipation and *GBA1* status (OR = 2.05, 95%CI = 1.18-3.64, P = 0.012), which is interesting as constipation has also been found to be predictive of cognitive decline in PD^28,29^. Finally, the frequency of hallucinations, which again have been shown to be a risk factor for dementia in PD^30^, was increased in *GBA1* mutation carriers (27.1 % vs 15.5%; ꭓ^2^ = 3.9, df = 1, 95%CI = -0.02-0.25, P = 0.049), albeit the association of *GBA1* mutations with hallucinations was not significant after correcting for confounders. Interestingly, when analysing the effect of *GBA1* variants by their severity^31^ (Supplementary Table 10), the association with decreased MoCA scores was only observed in mutations classified as “severe” (beta = -1.49, sd = 0.72, P = 0.039).

### Polygenic risk score analysis

Despite the significant enrichment of cases with early onset and/or family history of PD, who carry an increased *a priori* probability of a positive genetic finding, a monogenic cause for PD was not identified in 90.4% of families, of which 66.4% completed WGS and MLPA. A further 11.9% of cases carried a *GBA1* variant that significantly increases the risk of PD. We therefore wondered if other seemingly monogenic cases could be the result of increased risk of PD due to the cumulative effect of several risk variants, each contributing only a small fraction to the overall PD risk^32^. To answer this question, we calculated the PD polygenic risk score (PD-PRS) for each individual, but found that unit changes in the z-transformed PD-PRS were not positively or negatively associated with PD mutation status (Supplementary Figure 2a; OR = 1.07, 95%CI = 0.82-1.41, P = 0.624). Looking in more detail at the mutation-negative group, we found an association between the PD-PRS and a family history of PD specifically in cases with early onset (Supplementary Figure 2b; OR = 1.41, 95%CI = 1.02-1.94, P = 0.036), which suggests that a subset of mutation-negative early onset PD families might have pseudo-autosomal inheritance due to a shared increased load of common risk variants.

## DISCUSSION

The UK-based PFP study consists of early-onset and familial PD cases and their relatives, with a collection of detailed demographic, clinical, lifestyle, and environmental data, as well as biological samples for genetic testing. It aims to provide support for monogenic PD gene discovery while contributing to the characterisation of genotype-phenotype relationships of known monogenic forms of PD. The first phase of genetic screening for mutations in genes known to cause PD has been successfully completed for 718 families. Pathogenic causal mutations have been identified in 69 families, providing an overall diagnostic yield of 9.6% (13.8% in EOPD and 6% in fLOPD). This is in line with previous studies that found pathogenic mutations in known PD-related genes account for 5-10% of familial PD cases^33^.

Unsurprisingly, mutations in *LRRK2* were the most common cause of monogenic PD and were more frequent in the fLOPD group, although 16.7% of cases did not report a family history of PD and age of motor symptom onset ranged between 34 and 80 years. Age of symptom onset for *LRRK2* is reported to average 58–61 years, yet it frequently varies even within the same family^34^, probably reflecting the presence of disease-modifying genetic factors^35,36^. In addition, the seemingly sporadic nature of *LRRK2*-associated PD in many individuals is also likely due to its incomplete penetrance, which has been extensively described elsewhere^34,37,38^. While clinical characteristics are largely indistinguishable from idiopathic PD^34^, it has been suggested that *LRRK2*-associated PD has a milder phenotype and slower disease progression^39^. We found that *LRRK2* mutations were associated with an increased risk of dyskinesia and motor fluctuations compared to mutation-negative PD. This is in line with a large meta-analysis reporting an increased likelihood of motor complications in *LRRK2* G2019S carriers^40^. However, other studies comparing *LRRK2*-PD with idiopathic PD did not find an association between *LRRK2* status and incidence of dyskinesias^41,42^.

Biallelic mutations in *PRKN* were the second most frequently identified cause of monogenic PD and were present in 3.6% of families, all with EOPD. These individuals had an earlier age at symptom onset compared to mutation-negative PD cases, consistent with findings reported elsewhere^7,8,43^. We also observed lower MDS-UPDRS motor severity scores after adjusting for disease duration, indicating slower progression of motor symptoms compared to mutation-negative PD cases. In line with slower disease progression, there was significant association between biallelic *PRKN* carrier status and a decrease in the MDS-UPDRS part II scores, indicating reduced impact of motor symptoms on experiences of daily living. These findings are consistent with other studies, which have shown slower progression in biallelic *PRKN* carriers^7^. Previous studies have reported that postural symptoms^8^, dystonia, and psychiatric symptoms may be more common in *PRKN* carriers^7,44^, but we did not find evidence of this in our cohort.

In addition to monogenic PD-related genes, *GBA1* mutations were present in 10.2% of families, thus confirming *GBA1* as the most important genetic risk factor for PD. Family history of PD was present in most *GBA1* mutation carriers, often in a pattern akin to autosomal dominant inheritance. REM sleep behaviour disorder, a precursor of dementia in PD^25–27^, was more frequent in *GBA1* mutation carriers, as previously reported by others^45,46^. As expected, *GBA1* mutation carriers also performed worse in cognitive testing, in line with several studies showing worse cognitive outcomes in PD *GBA1* mutation carriers^47–52^. The detrimental effect of *GBA1* mutations on cognition was observed only in cases harbouring severe mutations (i.e., pathogenic mutations associated with neuronopathic forms of Gaucher disease), again corroborating previous studies^53^. However, it should be noted that other studies have found an association between the common risk variant E365K and cognitive decline in PD^46,51,54^.

In 90.4% of cases, no pathogenic mutations could be identified, which suggests that additional causative or contributing genetic factors are yet to be uncovered. It is possible that not all cases with early onset and/or familial PD have a monogenic form of the disease. We have found *GBA1* risk variants in 10.2% of our mutation-negative cohort, which increase the risk of PD in families that share *GBA1* risk variants. The incidence of *GBA1* mutations is significantly higher among PD patients, but the degree of pathogenicity and penetrance of different mutations is still debated^55^. Likewise, we have found a single heterozygous mutation in a recessive PD-related gene in another 1.9% of all index cases fully investigated with WGS and MLPA. These could represent truly monogenic PD, where the second mutation has yet to be identified due to technical constraints. Recently, long-read sequencing has identified complex structural variants in *PRKN* not detected by MLPA, including large inversions^56,57^. Conversely, there have been reports that heterozygous *PRKN* and *PINK1* carriers may have increased risk of developing PD symptoms with highly reduced penetrance^58–60^. However, other studies did not find an association between single heterozygous mutations in recessive PD-related genes and the risk of PD^61,62^. Interestingly, a recent study found that symptomatic heterozygous *PRKN* carriers had significantly reduced PRKN expression in peripheral blood mononuclear cells^63^. Furthermore, PRKN expression levels were decreased in symptomatic relative to asymptomatic family members carrying the same variants, suggesting the existence of additional genetic or epigenic mechanisms that regulate PRKN expression and could contribute to the risk of PD in monoallelic *PRKN* carriers^63^. Another possibility is that familial PD can be polygenic in nature, with relatives sharing multiple risk variants, each with a small risk effect, that increase the overall risk of PD among family members that share the same genetic background^32^. We did not find an association between the PD polygenic risk score and mutation status. However, in early-onset mutation-negative PD cases, an increasing PD-PRS was associated with familial status, which suggests that, at least in some families, a polygenic PD risk, compounded by the cumulative effect of many common risk variants, might contribute to a familial risk of PD, giving the appearance of pseudo-autosomal inheritance.

In addition to pathogenic mutations in well-established PD-related genes (*LRRK2*, *PRKN*, *PINK1*, *SNCA*, *PLA2G6* and *GBA1*), we identified pathogenic mutations in genes that have been reported to present as levodopa-responsive parkinsonism but typically present with alternative or atypical phenotypes. Mutations in *PNPLA6* cause Hereditary Spastic Paraplegia 39 (OMIM #612020), but levodopa-responsive parkinsonism has been reported in association with biallelic mutations, generally with additional clinical features^64,65^. Mutations in *SPG7* cause Hereditary Spastic Paraplegia 7 (OMIM #607259), which typically presents as pure spastic paraplegia but is often associated with complex phenotypes. Cases presenting with levodopa-responsive parkinsonism in association with biallelic *SPG7* mutations have been previously reported^66–68^. The *VCP* gene is typically associated with autosomal dominant Charcot-Marie Tooth type 2Y (OMIM #616687), frontotemporal dementia and/or amyotrophic lateral sclerosis 6 (OMIM #613954), or inclusion body myopathy with early-onset Paget disease and frontotemporal dementia (OMIM #167320). There are several reports of levodopa-responsive parkinsonism in association with pathogenic mutations in *VCP*^69–71^. Likewise, mutations in *GCH1* typically manifest as dopa-responsive dystonia (OMIM #128230), but several cases manifesting with autosomal dominant PD have been reported^72–74^. Finally, we found three individuals with fLOPD due to a pathogenic repeat expansion in *ATXN2*. Although typically manifesting as spinocerebellar ataxia 2 (OMIM #183090), *ATNX2* expanded CAG trinucleotide repeats have been identified in PD cases across multiple ancestries, most often in association with a family history of autosomal dominance^75–79^. Even though *ATXN2* repeat expansions have generally been considered a rather rare cause of PD^80^, they were the third most common cause of familial PD (and the second most common in late-onset disease) in this cohort.

Our study as some limitations. Over 90% of all recruited participants are of European ancestry, meaning that mutation rates cannot be generalised across populations. Further efforts are needed to recruit individuals from other ancestry groups. Despite our efforts to recruit family members, the number of recruited relatives is still relatively small. Several reasons account for this, namely, the fact that in adult-onset disorders such as PD, family members from older generations might no longer be available for study participation. In addition, the fact that this is a cross-sectional study without longitudinal follow-up might hamper recruitment of newly affected relatives at a future date. We cannot rule out a recruitment bias inherent to the study design, given the inability to recruit all eligible PD cases in a clinic-based study as compared to a community-based study.

In summary, we have identified a monogenic form of PD in 9.6% of recruited families. An additional 10.2% of families carried a *GBA1* variant. We have succeeded in building a cohort enriched for known pathogenic variants in PD-related genes, which will aid further characterization of genotype-phenotype associations, important for accurate diagnosis and prognosis prediction. The large number of families with a seemingly strong genetic component that remain without a molecular diagnosis presents an opportunity to uncover novel causative or high-risk conferring genetic variants and will be the focus of the next phase of the analysis. Currently, efforts are being made to recruit additional relatives from these unexplained families, in particular targeting families with a very early age at symptom onset or with multiple affected family members. As more samples are whole-genome sequenced from both affected and unaffected family members, segregation studies will be possible for demonstrating gene-disease associations, thereby facilitating new genetic discoveries. In addition, unaffected mutation carriers will allow for the examination of penetrance modifiers, thus providing insights into disease mechanisms and potential drug targets. PFP will continue to recruit from currently participating and new families until 2030.

## METHODS

### Subjects and clinical data collection

The PFP study has been reviewed and approved by the London Camden and King’s Cross Research Ethics Committee (REC – 15/LO/0097; IRAS ID – 162268) and is sponsored by the University College London Joint Research Office. The study is conducted in compliance with UK General Data Protection Regulation (GDPR) and the principles expressed in the Helsinki Declaration. PFP is registered with <u>www. clinicaltrials.gov</u> (NCT02760108). All participants provided written informed consent to study participation and data sharing. Participants could also opt to consent to confirmatory diagnostic genetic testing in case of a positive genetic finding, and to being re-contacted for further research studies, including therapeutic drug trials.

For this analysis, we included families recruited to PFP between 01/01/2015 and 24/02/2020, at 43 study sites across the UK (Figure 1). Eligible index cases had a clinical diagnosis of PD and met at least one of the following criteria: i) Motor symptom onset at or before the age of 45 (early onset PD); ii) At least one relative up to 3rd degree affected by PD (familial PD). We set the cut-off for early-onset disease at 45 years to specifically target individuals with higher *a priori* probability of recessive PD, given previous studies showing that the cumulative rate of pathogenic recessive mutations is considerably higher in younger age groups^8^. Whenever possible we also recruited affected and unaffected relatives of index cases. Participating individuals were at least 16 years old and had capacity to consent to participation. Participants were assessed only once during the study. For all participants, we collected demographic, environmental, medical, and family history data through questionnaires and a peripheral blood or saliva sample for DNA extraction. We also facilitated remote participation of participants who did not live near a study site. These participants completed shortened and simplified assessment booklets from home and donated samples through their local doctor. Patient questionnaires included: Parkinson’s Disease Quality of Life Questionnaire (PDQ-8), EQ-5D, Epworth Sleepiness Scale (ESS), REM Sleep Behavior Disorder Screening Questionnaire (RBDSQ), Hospital Anxiety and Depression Scale (HADS), Questionnaire for Impulsive-Compulsive Disorders in Parkinson’s Disease (QUIP), Fecal Incontinence and Constipation Questionnaire, Scales for Outcomes in Parkinson’s Disease - Autonomic (SCOPA-AUT), Parkinson’s Disease Sleep Scale (PDSS). Affected participants recruited on-site were also subject to a standardised structured interview and completed validated scales and questionnaires by experience raters to assess motor and non-motor symptoms, including: Montreal Cognitive Assessment (MoCA), Movement Disorder Society Unified Parkinson’s Disease Rating Scale (MDS-UPDRS), and the Modified Hoehn and Yahr Stages. Figure 1 shows an overview of the study protocol.

**Figure 1.**
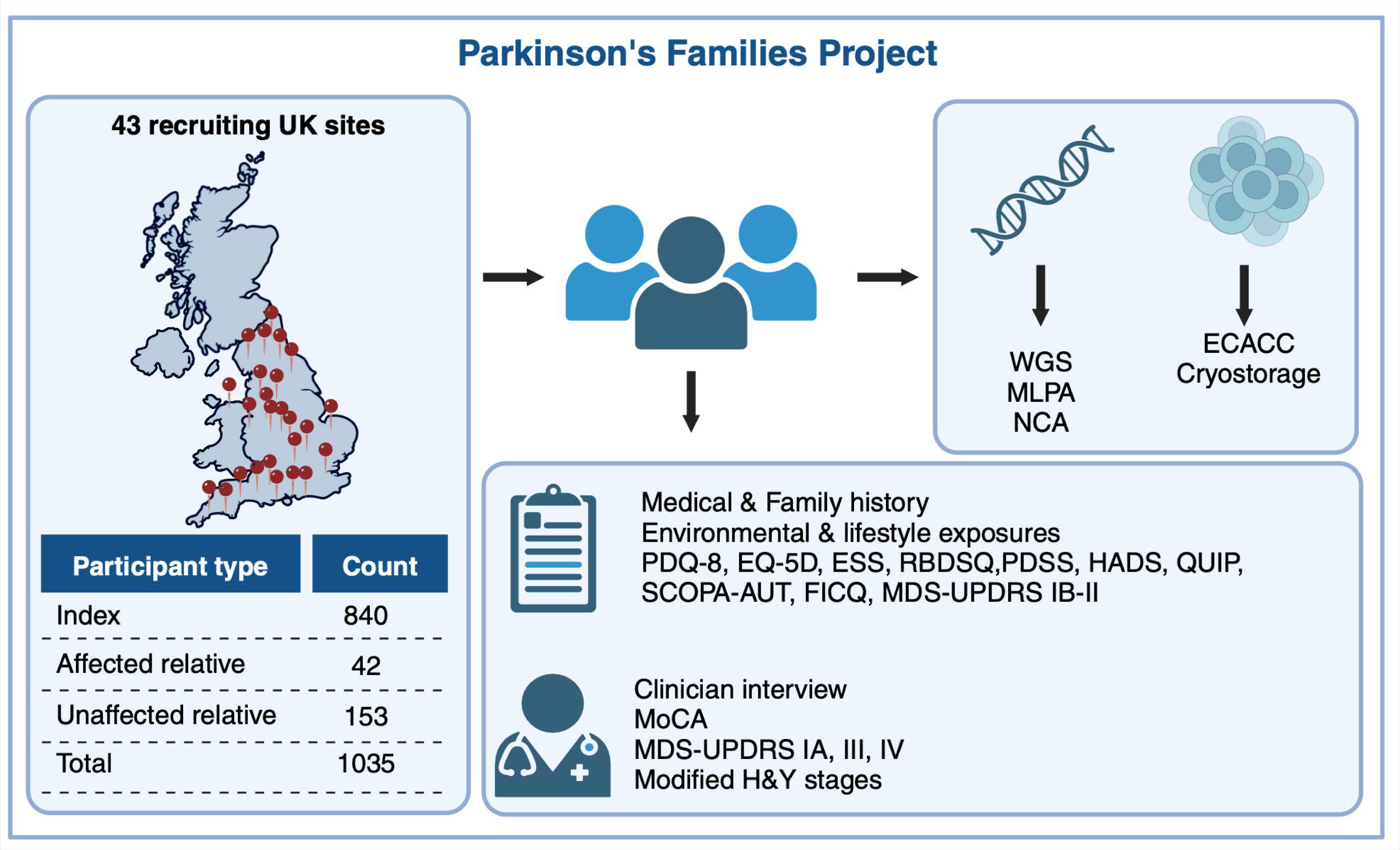
Parkinson’s Families Project overview. Participants are recruited across 43 sites in the UK. Index cases must be ≥18 years, have capacity to consent, have a diagnosis of PD with symptom onset ≤45 and/or family history of PD. All participants donate a blood sample for DNA extraction. Affected participants additionally donate blood for peripheral blood lymphocyte extraction, which are sent to the European Collection of Authenticated Cell Cultures (ECACC) for transformation into lymphoblastoid cell lines and storage. All affected participants fill out a questionnaire with detailed medical and family history, environmental, drug and lifestyle exposures, as well as the following questionnaires: Parkinson’s Disease Questionnaire (PDQ-8), EQ-5D, Epworth Sleepiness Scale (ESS), REM Sleep Behavior Disorder Screening Questionnaire (RBDSQ), Panic Disorder Severity Scale (PDSS), Hospital Anxiety and Depression Scale (HADS), Questionnaire for Impulsive-Compulsive Disorders in Parkinson’s Disease (QUIP), Scales for Outcomes in Parkinson’s disease - Autonomic Dysfunction (SCOPA-AUT), Fecal Incontinence and Constipation Questionnaire (FICQ), MDS-UPDRS parts IB and II. Affected participants recruited on-site are also assessed by an experienced investigator, who rates the MDS-UPDRS parts IA, III and IV, MoCA and Hoehn & Yahr scales. WGS: whole-genome sequencing; MLPA: multiplex ligation-dependent probe amplification assay; NCA: NeuroChip genotyping array.

Participants with partially completed MDS-UPDRS ratings that fell below the threshold defined by Goetz and colleagues were excluded from downstream analyses^81^. Subjects were classified into motor subtypes (tremor dominant [TD], postural instability and gait difficulty [PIGD] or intermediate) based on the methodology defined by Stebbins and colleagues^82^. If items required for classification were missing, individuals were labelled as “unclassifiable”. To account for differences in disease duration at assessment, we computed a motor severity score that consists of the ratio between the total MDS-UPDRS part III score and disease duration from reported symptom onset. Based on the MDS-UPDRS part IV, we also computed composite scores for dyskinesia (sum of items 4.1 and 4.2) and motor fluctuations (sum of items 4.3-4.5). Items of the MDS-UPDRS were categorised as present if the composite score was ≥ 1, except depression (item 1.3) and apathy (item 1.4), which were considered present only if sustained over more than one day at a time (score ≥ 2). REM sleep behaviour disorder was considered present if the RBDSQ was >5.

### Clinical data storage and management

Data collected is held on REDCap® (Research Electronic Data Capture), a secure web-based Hypertext Preprocessor (PHP) software with a MySQL database back-end (https://www.project-redcap.org). It is tried and tested for use in managing clinical studies and trials, longitudinal studies and surveys^83^. The web host, network connection and storage is Information Governance Toolkit (IGT)-compliant and ISO27001-certified, according to data security best practices. Personally identifiable information is held in a database that is separated from the main study database. Members of the study team at each site only have access to records for participants recruited at their site. The databases will be maintained until 2034 for genetic/epidemiological research, under the custodianship of Prof. Huw Morris to enable the long-term follow-up of patients recruited to this study. All clinical data were processed, stored, and disposed in accordance with all applicable legal and regulatory requirements, including the Data Protection Act 1998 and any amendments thereto.

### Sample collection and storage

DNA was extracted from EDTA blood or saliva samples (saliva collection kit: Oragene® OG-500, DNA Genotek Inc.) by LGC Biosearch Technologies™. DNA is stored in secure freezers at University College London. Affected participants additionally donated ACD blood that was sent to the European Collection of Authenticated Cell Cultures (ECCAC, https://www.culturecollections.org.uk/collections/ecacc.aspx), in Wiltshire, UK, for peripheral blood lymphocytes (PBLs) extraction and transformation into lymphoblastoid cell lines. These cell lines provide an ongoing source of DNA for future studies, and may be used for disease models or the generation of induced pluripotent cell lines. Cell lines are stored at the ECACC encoded by the unique PFP study identifier.

### Genetic analysis

#### Whole Genome Sequencing (WGS)

DNA samples from 585 participants were sequenced within the Global Parkinson’s Genetics Program (GP2) Monogenic Network^84,85^. Briefly, samples were sequenced with Illumina short-read WGS at Psomagen, with a mean coverage of 30x. 150 bp paired-end reads were aligned to the human reference genome (GRCh38 build) using the functional equivalence pipeline^86^. Sample processing and variant calling were performed using DeepVariant v.1.6.1^87^. Joint-genotyping was performed using GLnexus v1.4.3 with the preset DeepVariant WGS configuration^88^. Samples were retained for downstream analyses after passing the quality control with the quality metrics defined by the Accelerating Medicines Partnership Parkinson’s Disease program (AMP-PD; https://amp-pd.org)^89^. Variant annotation was performed with Ensembl Variant Effect Predictor^90^. A target list of *GBA1* variants were called using the Gauchian v.1.0.2 tool (https://github.com/Illumina/Gauchian)^91^. The length of STRs in *ATXN2* and *ATXN3* was estimated in whole-genome sequence data using the ExpansionHunter v.5.0.0 software^92^. All the pipelines used are available on GitHub (https://github.com/GP2code/GP2-WorkingGroups/tree/main/MN-DAWG-Monogenic-Data-Analysis). Additional details on variant interpretation are available in Supplementary Materials. A further 39 participants were analysed with WGS as part of the 100,000 Genomes Project^93^.

#### Next-Generation Targeted Sequencing (NGS)

DNA samples of an additional three participants underwent diagnostic genetic screening using next-generation sequencing (Illumina MiSeq or HiSeq) of a panel of seven genes (*FBXO7*, *LRRK2*, *PRKN*, *PARK7*, *PINK1*, *SNCA*, *VPS35*) and MLPA gene dosage analysis of three genes (*PRKN*, *PINK1*, *SNCA*), as described in the next section. Pathogenic or likely pathogenic variants were confirmed with bi-directional Sanger sequencing.

#### Multiplex Ligation-dependent Probe Amplification (MLPA)

Samples from 827 participants were screened for copy number variants (CNVs) using the SALSA MLPA EK5-FAM reagent kit and the SALSA MLPA Probemix P051, according to the manufacturer’s instructions (MRC-Holland, Amsterdam, The Netherlands). Where DNA was available, we additionally screened relatives of index cases with a CNV. PCR fragments were analysed by capillary electrophoresis using an ABI 3730XL genetic analyzer (Applied Biosystems). Data was analysed using the Coffalyser.Net™ (MRC-Holland) or GeneMarker® (SoftGenetics®, PA, USA) software packages, according to the supplied protocols.

#### SNP Array Genotyping

Quantity and purity of DNA were determined with a Qubit fluorometric assay (Invitrogen) and a NanoDrop spectrophotometer (Thermo Fisher Scientific, UK), respectively. Samples were diluted to a standard concentration in molecular grade nuclease-free water (Thermo Fisher Scientific, UK). We genotyped 849 DNA samples from 698 families using the Illumina NeuroChip array (NCA), which consists of a 306,670 SNP backbone (Infinium HumanCore-24 v1.0) with added custom content covering 179,467 neurodegenerative disease-related variants^94^. We manually clustered the genotypes using Illumina GenomeStudio v2.0 (Illumina Inc., San Diego, CA, USA), based on the protocol by Guo and colleagues^95^. We curated a list of *GBA1* PD risk variants and GD-causing mutations, as well as pathogenic and likely pathogenic SNVs and indels from 10 PD causing genes (*PRKN, DJ-1, PINK1, ATP13A2, FBXO7, SCNA, LRRK2, VCP, VPS35, DCTN1*), from ClinVar (https://www.ncbi.nlm.nih.gov/clinvar/, accessed on the 18/01/2023)^96^. We added any additional variants from PD-related genes classified as definitely pathogenic in the MDSGene database (https://www.mdsgene.org/, accessed on the 21/02/2023)^97^. Probes for 131 of these variants were present in the Neurochip array and were systematically screened for in all index cases using a custom R script (Supplementary Table 2). We evaluated the accuracy of the NCA probes of interest by comparing their performance against other methods, as described in Supplementary Materials.

For additional downstream analyses, we performed standard quality control in PLINK v1.9^98^. Briefly, we excluded samples with genotype missingness >5% (which can indicate poor quality of DNA sample), mismatch between clinical and genetically determined sex (which could be due to a sample mix-up), and excess heterozygosity defined as individuals who deviate > 3SD from the mean heterozygosity rate (which can indicate sample contamination)^99^. We excluded variants if the call rate was <95%. Pairwise identity-by-descent (IBD) analysis was performed to infer relatedness across all samples and identify cryptic familial relationships using the KING tool (https://www.kingrelatedness.com/)^100^. Ancestry was genetically determined using GenoTools (https://github.com/dvitale199/GenoTools)^101,102^. To perform polygenic risk score analysis, genotypes were imputed against the TOPMed reference panel (version R2; https://www.nhlbiwgs.org/) using the TOPMed Imputation Server (https://imputation.biodatacatalyst.nhlbi.nih.gov) using Minimac4 (version 1.7.3)^103^. Imputed variants were excluded if the imputation info R^2^ score was ≤ 0.3. Following imputation, variants with missingness > 5% and minor allele frequencies < 1% were also excluded. Polygenic risk scores were computed in PRSice-2 (https://choishingwan.github.io/PRSice/)^104^ based on summary statistics from the largest Parkinson’s disease case-control genome-wide association study (GWAS) to date^105^.

### Statistical analyses

For statistical analysis, we classified PD cases into the following categories: i) Sporadic early-onset PD (sEOPD): motor symptom onset ≤ 45 years, no family history of PD; ii) Familial early-onset PD (fEOPD): motor symptom onset ≤ 45 years, positive family history of PD; iii) Familial late-onset PD (fLOPD): motor symptom onset > 45 years, positive family history of PD. *GBA1* variants were classified by severity according to the GBA1-PD browser (https://pdgenetics.shinyapps.io/gba1browser/, accessed on the 25^th^ May 2024)^31^. For statistical purposes, the mutation-negative and monoallelic *PRKN* mutation groups comprise only individuals fully investigated with WGS and MLPS, to ensure that no undetected mutations are present. Likewise, the *GBA1* mutation group excludes individuals not investigated with WGS. We compared demographic and clinical features using Mann-Whitney U-test for continuous variables and Fisher’s exact tests or Chi-squared tests for proportions. We investigated the effect of the *LRRK2*, *PRKN* and *GBA1* genetic status on clinical features using linear regression for continuous scores or logistic regression for categorical scores, adjusting for sex, age at assessment, and disease duration at assessment, where appropriate. We used multinomial logistic regression to analyse motor subtype, using the tremor dominant group as the reference. For analysis of the modified Hoehn & Yahr stages, we used the 0-1.5 group as the reference. For the polygenic risk score analysis, scores were z-transformed and used in logistic regression models to predict the dependent variables. All p-values are two-tailed. We used R version 4.2.1 to perform statistical analyses^106^.

## Data Availability

The data, code, protocols, and key lab materials used and generated in this study are listed in a Key Resource Table alongside their persistent identifiers at 10.5281/zenodo.12549398.

## ACKNOWLEDGEMENTS

PFP has received support from the Janet Owen’s bequest fund, the Walker-Peltz charitable fund, the Medical Research Council (MRC-G1100643), Cure Parkinson’s Trust, Parkinson’s UK (K-1501), and the National Institute for Health Research (NIHR) Clinical Research Network (CRN) North Thames. The funders played no role in study design, data collection, analysis and interpretation of data, or the writing of this manuscript. A full list of PFP Study Group members is available in Supplementary Materials.

This research was funded in part by Aligning Science Across Parkinson’s [Grant number: ASAP-000478] through the Michael J. Fox Foundation for Parkinson’s Research (MJFF). For the purpose of open access, the author has applied a CC BY public copyright licence to all Author Accepted Manuscripts arising from this submission.

Part of the whole-genome sequencing data used in the preparation of this article were obtained from Global Parkinson’s Genetics Program (GP2). GP2 is funded by the Aligning Science Across Parkinson’s (ASAP) initiative and implemented by The Michael J. Fox Foundation for Parkinson’s Research (https://gp2.org/). For a complete list of GP2 members see https://gp2.org/.

This research was in part made possible through access to data in the National Genomic Research Library, which is managed by Genomics England Limited (a wholly owned company of the Department of Health and Social Care). The National Genomic Research Library holds data provided by patients and collected by the NHS as part of their care and data collected as part of their participation in research. The National Genomic Research Library is funded by the National Institute for Health Research and NHS England. The Wellcome Trust, Cancer Research UK and the Medical Research Council have also funded research infrastructure.

The authors would like to thank study participants and referring clinicians, without whom this study would not be possible. Figures created with BioRender.com.

## DATA AVAILABILITY

A pseudo-anonymised cleaned dataset is available from https://doi.org/10.5281/zenodo.12549399. The data, code, protocols, and key lab materials used and generated in this study are listed in a Key Resource Table alongside their persistent identifiers at <u>10.5281/zenodo.12549398.</u>

Array data has been deposited at the European Genome-phenome Archive (EGA), which is hosted by the EBI and the CRG, under accession number EGAS00001007906. Further information about EGA can be found on https://ega-archive.org and “The European Genome-phenome Archive in 2021” (https://doi.org/10.1093/nar/gkab1059).

For whole-genome sequence data obtained from the 100,000 Genomes Project, research on the de-identified patient data used in this publication can be carried out in the Genomics England Research Environment subject to a collaborative agreement that adheres to patient led governance. All interested readers will be able to access the data in the same manner that the authors accessed the data. For more information about accessing the data, interested readers may contact or access the relevant information on the Genomics England website: https://www.genomicsengland.co.uk/research.

Data (DOI 10.5281/zenodo.10962119, release 7) used in the preparation of this article were partially obtained from the Global Parkinson’s Genetics Program (GP2). To obtain access to de-identified individual level data, interested readers must register to access the AMP PD Knowledge Platform: https://amp-pd.org/researchers/data-use-agreement.

## CODE AVAILABILITY

Raw SNP array data was clustered in GenomeStudio v2.0 (RRID:SCR_010973) according to the protocol described by Guo et al. (ref.^95^) and quality control performed in Plink v1.9 (RRID:SCR_001757). Genetic ancestry was determined using Genotools (doi: 10.5281/zenodo.10443258). Sample relatedness was inferred using KING (RRID:SCR_009251). Polygenic risk scores were computed in PRSice-2 (RRID:SCR_017057). WGS processing, quality control, joint genotyping and variant calling of data generated in Genomic England in the 100,000 Genomes Project was done according to the protocol defined in ref.^93^. WGS processing, quality control, joint genotyping and variant calling of data generated in GP2 was performed using DeepVariant v.1.6.1 (https://github.com/google/deepvariant) and GLnexus v1.4.3 (https://github.com/dnanexus-rnd/GLnexus) according to pipelines available at https://github.com/GP2code. Variants were annotated with Ensembl Variant Effect Predictor (RRID:SCR_007931). GBA1 variants were called using Gauchian v.1.0.2 (https://github.com/Illumina/Gauchian). Short tandem repeat sizing was performed using ExpansionHunter v.5.0.0 (https://github.com/Illumina/ExpansionHunter). For data generated by fragment analysis, GeneMapper® v5.0 (RRID:SCR_014290) was used. MLPA data was analysed using GeneMarker® (RRID:SCR_015661) or Coffalyser.Net (freely available from https://www.mrcholland.com/technology/software). Statistical analyses were performed in R v4.2.1 (RRID:SCR_001905) using basic statistical packages (stats v4.2.1, nnet v7.3.19). Other packages used include dplyr (v1.1.4), tidyr (v1.3.0), ggplot2 (v3.4.4), data.table (v1.14.8), broom (v1.0.5), purrr (v1.0.2), knitr (v1.45), forcats (v1.0.0) and plinkQC (v0.3.4).

## AUTHOR CONTRIBUTIONS

RR, CT, MMXT and HRM designed the study. MMXT, LW and RR prepared samples for SNP array genotyping. RR performed clustering, quality control and analysis of SNP array data. EJS prepared samples for WGS. MMXT, CT and RR performed and analysed MLPA experiments. RR performed and analysed *ATXN2* and *ATXN3* fragment analysis experiments. ZHF performed WGS variant calling and annotation of GP2-generated WGS data, and developed the pipeline for STR expansion calling. RR performed variant interpretation of GP2-generated WGS data. ZHF and RR performed STR expansion analysis on GP2 and Genomics England WGS data, respectively. JH, RL and JP performed and interpreted diagnostic targeted sequencing data. MH, MMXT, MP, RR, RT, SJ and TMS collected and processed clinical data. RR performed statistical analysis and interpreted the data. ABS and CB are the leads of the Global Parkinson’s Genetics Program. CK, LML and ZHF are members of the GP2 Monogenic Network, of which CK is the lead. RR, HRM, PRJ, HH and NWW are the leads of the Parkinson’s Families Project. KPB and AHVS advised on the project. CT, RR and TMS wrote the initial draft of the manuscript. All authors read and approved the final manuscript.

