## Supplementary Materials for "Parkinson’s Families Project: a UK-wide study of early onset and familial Parkinson’s disease"

### Supplementary Methods

#### Whole genome sequencing variant interpretation

In whole genome sequencing (WGS) data generated in the 100,000 Genomes Project, the "Parkinson Disease and Complex Parkinsonism" gene panel from Genomics England PanelApp (https://panelapp.genomicsengland.co.uk/) was used for gene prioritisation. Data processing, variant calling and variant interpretation were performed by Genomics England[1]. In data generated in the GP2 Monogenic Network, a curated panel of 74 genes (Supplementary Table 11) selected on the basis of association with PD or Parkinsonism was screened for potentially disease-causing variants. High-quality variants with a call rate >0.95, genotype quality (GQ) >20, read depth (DP) >5, and heterozygous allele balance (AB) between 0.2 and 0.8 were retained. Dominantly inherited protein coding SNVs/indels and splice site variants with the maximum credible genetic ancestry group allele frequency >0.005 were excluded[2], as well as homozygous or potential compound heterozygous variants with >1 homozygote for the alternative allele in gnomAD v4.1.0 genomes (https://gnomad.broadinstitute.org)[3]. In the first instance, we used Clinvar annotations to define pathogenicity. Cases were considered solved if variants classified as "Pathogenic" or "Likely Pathogenic" were present and the zygosity fit the known mode of inheritance for each gene. Novel variants and variants of unknown significance (VUS) were subjected to further variant interpretation according to the ACMG classification and the subsequent ACGS guidelines for variant classification in rare disease[4,5]. We used online tools such as Franklin (https://franklin.genoox.com/- Franklin by Genoox) and Varsome (https://varsome.com/) to assist in variant interpretation[6]. We also performed a literature search for additional information on the interrogated variants. When available, we incorporated segregation data into the decision tree. After this process, variants still classified as "unknown significance" were excluded. We use the term "compound heterozygosity" to define the presence of two pathogenic variants in recessive genes, even in cases where it was not possible to confirm that the variants were in *trans* due to lack of DNA from the parents. In cases where two pathogenic mutations were in very close proximity in the genome, we used the desktop application of Integrative Genomics Viewer (v2.17.4; https://igv.org/) to confirm if the variants were in the same allele (*cis*) or in separate alleles (*trans*)[7]. An example can be seen in Supplementary Figure 3.

#### Validation of *ATXN2* and *ATXN3* short tandem repeats (STR)

For a subset of cases with STR calling from WGS, we additionally performed molecular analysis of short tandem repeats (*n*=33 for *ATXN2* and *n*=34 for *ATXN3*). Identification of repeat expansions in *ATXN2* and *ATXN3* genes was performed as previously described[8]. Briefly, amplification of the CAG trinucleotide repeat region was performed using fluorescent 6FAM-labelled primers (see Supplementary Table 12 for primer sequences). The amplified PCR fragments were sized by capillary electrophoresis on an ABI3730XL sequencer (Applied Biosystems) and the electropherogram peaks analyzed using GeneMapper® v5.0 software. Pathogenic expansions were defined using consensus thresholds from the literature[9]. Results from fragment size analysis were used to validate the performance of the ExpansionHunter algorithm in whole genome sequencing data (Supplementary Figure 4).

**Quantification of NeuroChip probe performance**

In a subset of 537 index cases, both NeuroChip genotyping array (NCA) and whole genome and/or targeted sequencing were available. We calculated the proportion of false positive and false negative genotype calls of all disease-causing mutations detected by NCA, considering whole genome or targeted sequencing as the reference (Supplementary Table 2). Of the 19 unique mutations detected using the NCA, there was a single probe that resulted in a false positive genotype (*GBA1* p.Ser235Pro). We then inspected the SNP graph for this variant in GenomeStudio$^{TM}$, which confirmed a poorly clustered SNP. Given its low sensitivity, we ignored results from this probe. Additionally, one of the probes (*GBA1* p.Arg159Trp) resulted in a false negative result. NCA-determined genotypes matched with sequencing data in 95.2% (40/42) of cases, indicating a very high sensitivity for most probes under assessment. However, given the limited number of probes included in the NCA array, its overall sensitivity to detect a pathogenic variant was 34.2%.

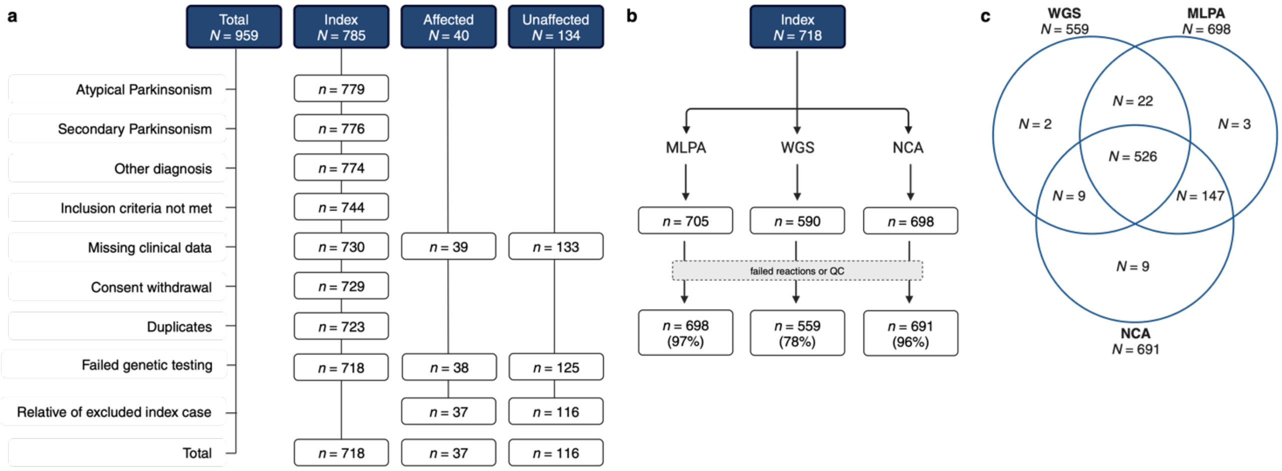

**Supplementary Figure 1. Final participant selection and investigations. a.** Flow chart of participant exclusions by participant type. **b.** Flow chart of investigations performed on index cases. **c.** Venn diagram of the overlap between each testing method in index cases. Targeted sequencing of a PD gene panel was available for an additional three index cases who did not have WGS. MLPA: multiplex ligation-dependent probe amplification assay; WGS: whole-genome sequencing; NCA: NeuroChip genotyping array.

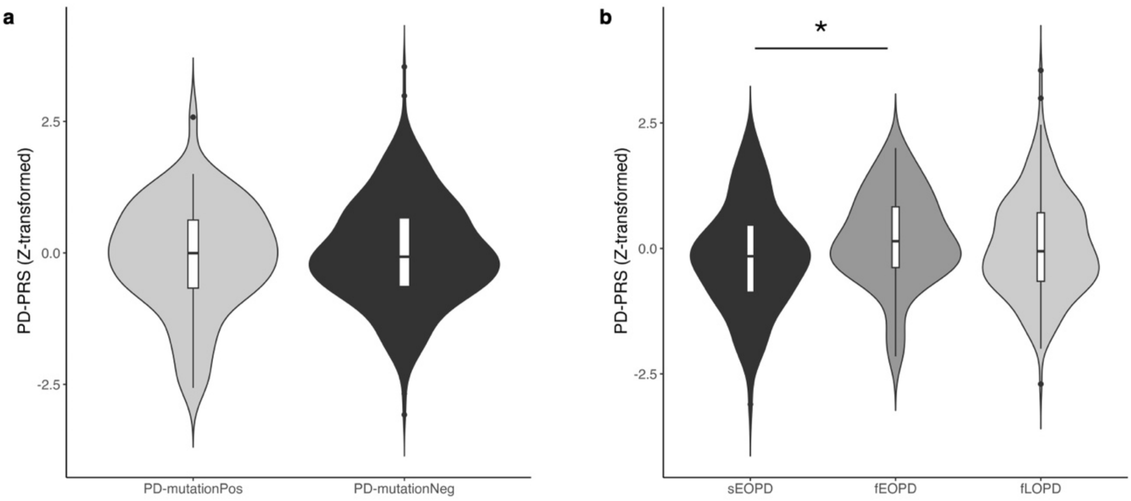

**Supplementary Figure 2. Violin plots of Z-transformed PD polygenic risk scores (PD-PRS). a.** Monogenic (*n*=67) vs mutation-negative (*n*=418) PD. Logistic regression of z-PRS scores against mutation status, with age at onset, sex, and PC1-5 added as covariates **b.** Mutation-negative sEOPD (*n*=121) vs fEOPD (*n*=60) vs fLOPD (*n*=237). Multinomial regression of z-PRS scores against disease category, with sex and PC1-5 added as covariates. Mutation negative cases exclude pathogenic or risk *GBA1* variant carriers, single heterozygous carriers of pathogenic variants in *PINK1* and *PRKN*, and cases that did not complete both WGS and MLPA. NCA data was used to compute the PD-PRS based on PD case-control GWAS summary statistics from Nalls *et al*. Statistical analysis was performed using logistic or multinomial regression, as appropriate. Box plots with median (centre line), upper and lower quartiles (box limits), 1.5x interquartile range (whiskers) and outliers (points) represented. *, $P<0.05$.

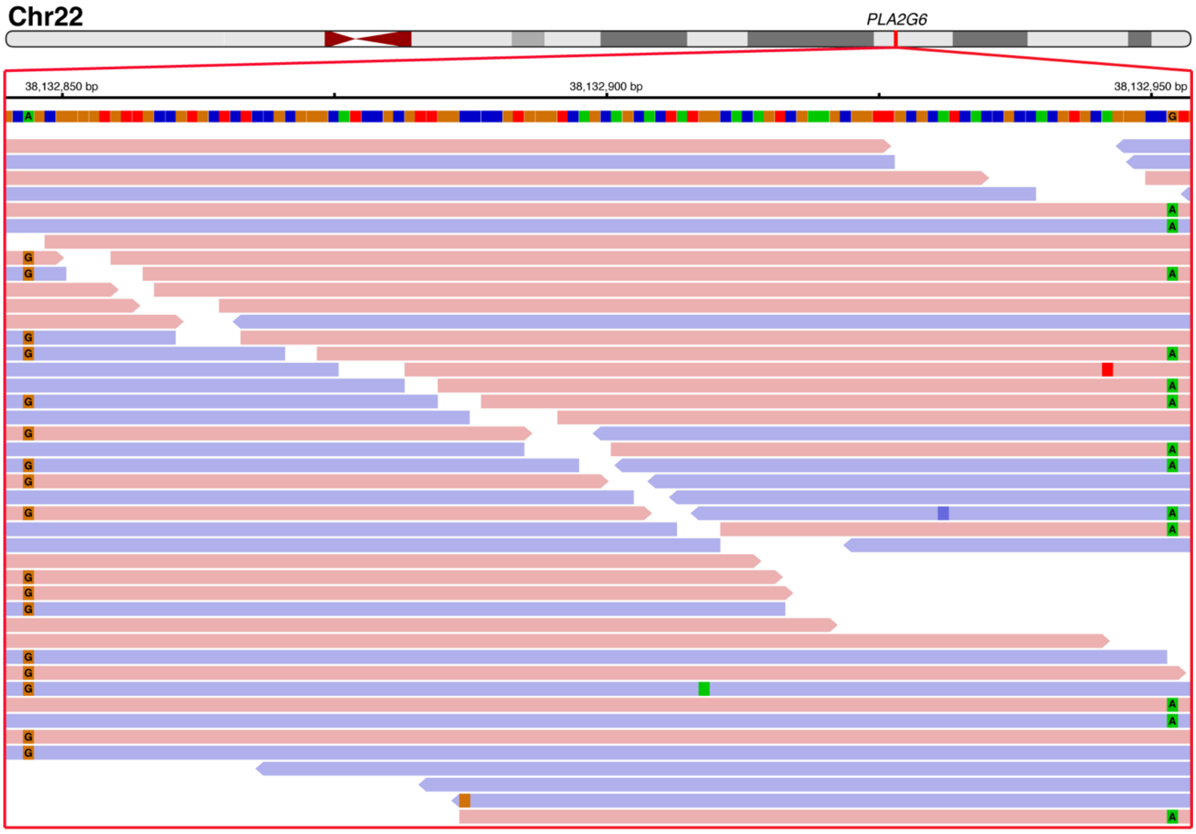

**Supplementary Figure 3. Integrative Genomics Viewer window of a 105 bp region in *PLA2G2*.** Two missense pathogenic variants were detected at positions c.1061T>C and c.956C>T. Given the close proximity between the variants, the possibility that the two variants might be segregating together in an LD block was interrogated. In reads that span the entire 105 bp region, it is apparent that the variants are never present in the same read, confirming that they are not part of a haplotype and were inherited independently from each parent.

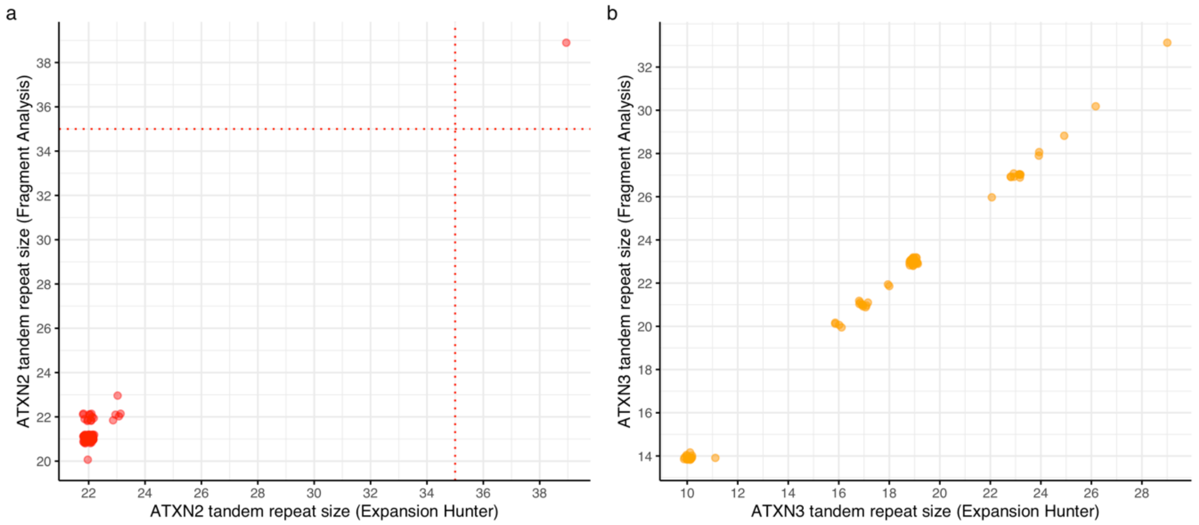

**Supplementary Figure 4*. Correlation of short tandem repeat (STR) size determined by fragment analysis (FA) and Expansion Hunter (EH). a.*** *ATXN2* STR length in 33 individuals (66 alleles) was 21.5 ± 2.2 for FA and 22.3 ± 2.1 for EH. Spearman correlation coefficient ($\rho$) = 0.548, P-value = 1.94e-06. The red doted lines indicate the threshold for pathogenicity (≥ 35 repeats). **b.** *ATNX3* STR length in 34 individuals (68 alleles) was 21.6 ± 4.8 for FA and 17.6 ± 4.8 for EH ($\rho$ = 0.999, P-value = < 2.2e-16). No alleles reached the threshold for pathogenicity (≥ 56 repeats), which is therefore not represented in the plot.

**Supplementary Table 1.** Variant details of families carrying PD-causing pathogenic mutations

| Family ID | Diagnosis | Group | Zygosity | Gene | Variant 1 | Variant 2 | Risk variants | Investigations |
|---|---|---|---|---|---|---|---|---|
| 1 | PD | fLOPD | HET | *LRRK2* | G2019S | | | NCA, MLPA |
| 2 * | PD | fLOPD | HET | *LRRK2* | G2019S | | | NCA, MLPA |
| 3 | PD | sEOPD | HET | *LRRK2* | G2019S | | | NCA |
| 4 | PD | sEOPD | HET | *LRRK2* | G2019S | | | NCA |
| 5 | PD | fLOPD | HET | *LRRK2* | G2019S | | | NCA, MLPA |
| 6 | PD | fLOPD | HET | *LRRK2* | G2019S | | | NCA, MLPA |
| 7 | PD | sEOPD | HET | *LRRK2* | G2019S | | | NCA, WGS |
| 8 ^ | PD | fLOPD | HET | *LRRK2* | G2019S | | | NCA, MLPA |
| 9 ^ | PD | fLOPD | HET | *LRRK2* | G2019S | | | NCA, MLPA |
| 10 | PD | fLOPD | HET | *LRRK2* | G2019S | | | NCA, MLPA |
| 11 | PD | fLOPD | HET | *LRRK2* | G2019S | | | NCA, MLPA |
| 12 | PD | fEOPD | HET | *LRRK2* | G2019S | | | NCA, MLPA |
| 13 | PD | sEOPD | HET | *LRRK2* | G2019S | | | NCA |
| 14 | PD | sEOPD | HET | *LRRK2* | G2019S | | | NCA, WGS |
| 15 | PD | fLOPD | HET | *LRRK2* | G2019S | | *GBA1* L29Afs*18 | NCA, MLPA, WGS |
| 16 | PD | fLOPD | HET | *LRRK2* | G2019S | | | NCA, MLPA |
| 17 | PD | fEOPD | HET | *LRRK2* | G2019S | | | NCA, MLPA |
| 18 | PD | fLOPD | HET | *LRRK2* | G2019S | | | NCA, MLPA |
| 19 | PD | fLOPD | HET | *LRRK2* | G2019S | | | NCA, MLPA |
| | PD | fLOPD | HET | *LRRK2* | G2019S | | | NCA, MLPA |
| 20 | PD | fLOPD | HET | *LRRK2* | G2019S | | | NCA, MLPA, WGS |
| 21 | PD | fLOPD | HET | *LRRK2* | G2019S | | | NCA, MLPA |
| 22 | PD | fLOPD | HET | *LRRK2* | G2019S | | | NCA, MLPA |
| 23 | PD | fLOPD | HET | *LRRK2* | G2019S | | | NCA, MLPA, WGS |
| | Unaffected | NA | HET | *LRRK2* | G2019S | | | NCA, MLPA |
| | Unaffected | NA | - | - | - | | | NCA, MLPA |
| 24 | PD | fLOPD | HET | *LRRK2* | G2019S | | | NCA, MLPA |
| 25 | PD | fLOPD | HET | *LRRK2* | G2019S | | | NCA, MLPA |
| 26 | PD | fLOPD | HET | *LRRK2* | G2019S | | | NCA, MLPA |
| 27 | PD | fLOPD | HET | *LRRK2* | G2019S | | | NCA, MLPA, WGS |
| 28 | PD | fLOPD | HET | *LRRK2* | G2019S | | | MLPA, WGS |
| 29 | PD | fLOPD | HET | *LRRK2* | R1441C | | | NCA, MLPA, WGS |
| 30 | PD | fLOPD | HET | *LRRK2* | N1437S | | | NCA, MLPA, WGS |
| 31 | PD | fLOPD | HET | *ATXN2* | (CAG)37 | | | NCA, MLPA, WGS |
| | PD | fLOPD | - | - | - | | | NCA, MLPA |
| 32 | PD | fLOPD | HET | *ATXN2* | (CAG)39 | | | NCA, MLPA, WGS |
| 33 | PD | fLOPD | HET | *ATXN2* | (CAG)37 | | | NCA, MLPA, WGS |
| 34 | PD | fLOPD | HET | RAB32 | Ser71Arg | | | NCA, MLPA, WGS |
| 35 | PD | sEOPD | HOM | *PNPLA6* | P1297S | P1297S | | NCA, MLPA, WGS |
| 36 | PD | fEOPD | COMP_HET | *SPG7* | N288* | A510V | | NCA, MLPA, WGS |
| 37 | Dystonia-Parkinsonism | sEOPD | COMP_HET | *PLA2G6* | T319M | L354P | | NCA, MLPA, WGS |
| 38 | PD | fEOPD | COMP_HET | *PRKN* | C166Y | Exon 1-3 deletion | | NCA, MLPA, WGS |
| 39 | PD | fEOPD | COMP HET | *PRKN* | Exon 3-4 deletion | C253Y | | NCA, MLPA, WGS |
| | PD | fEOPD | COMP HET | *PRKN* | Exon 3-4 deletion | C253Y | | NCA, MLPA, NGS gene panel |
| | PD | fEOPD | COMP HET | *PRKN* | Exon 3-4 deletion | C253Y | | NCA, MLPA, WGS |

**Supplementary Table 1.** Variant details of families carrying PD-causing pathogenic mutations

| Family ID | Diagnosis | Group | Zygosity | Gene | Variant 1 | Variant 2 | Risk variants | Investigations |
|---|---|---|---|---|---|---|---|---|
| | Unaffected | NA | - | - | - | - | | NCA, MLPA |
| | Unaffected | NA | HET | - | Exon 3-4 deletion | - | | NCA, MLPA |
| 40 | PD | sEOPD | HOM | *PRKN* | Q34Rfs* | Q34Rfs* | | NCA, MLPA, WGS |
| 41 | PD | sEOPD | COMP_HET | *PRKN* | G284R | C166Hfs*18 | *GBA1* T408M | NCA, WGS |
| 42 | PD | sEOPD | COMP_HET | *PRKN* | G430D | P113fs*51 | | NCA, MLPA, WGS |
| 43 | PD | sEOPD | COMP_HET | *PRKN* | N52Mfs*59 | W74Cfs*8 | | NCA, MLPA, NGS gene panel |
| 44 * | PD | sEOPD | COMP_HET | *PRKN* | P113fs*51 | Exon 2 deletion | | NCA, MLPA, WGS |
| 45 | PD | sEOPD | HOM | *PRKN* | R275W | R275W | | NCA, MLPA |
| | Unaffected | NA | HET | *PRKN* | R275W | - | | NCA, MLPA |
| 46 | PD | fEOPD | COMP HET | *PRKN* | G430D | R275W | | NCA, MLPA |
| | Unaffected | NA | HET | *PRKN* | G430D | - | | NCA, MLPA |
| 47 | PD | sEOPD | COMP HET | *PRKN* | R275W | P113fs*51 | | NCA, MLPA, WGS |
| | Unaffected | NA | HET | *PRKN* | R275W | - | | NCA, MLPA |
| | Unaffected | NA | HET | *PRKN* | Exon 3 deletion † | - | | NCA, MLPA |
| 48 | PD | fEOPD | COMP_HET | *PRKN* | R275W | Exon 4 deletion | | NCA, MLPA, WGS |
| 49 | PD | sEOPD | COMP_HET | *PRKN* | R275W | Exon 4 deletion | | NCA, MLPA, WGS |
| 50 | PD | fEOPD | COMP_HET | *PRKN* | R275W | Exon 5 deletion | | NCA, MLPA, WGS |
| 51 | PD | fEOPD | COMP_HET | *PRKN* | R275W | Exon 8 deletion | | NCA, MLPA, WGS |
| 52 | PD | sEOPD | HOM | *PRKN* | R275W | R275W | | NCA, MLPA |
| 53 | PD | fEOPD | COMP_HET | *PRKN* | R42P | Exon 3 duplication | | NCA, MLPA, WGS |
| 54 | PD | sEOPD | COMP_HET | *PRKN* | T240M & P437L | Exon 2 deletion | | NCA, MLPA, WGS |
| 55 | PD | sEOPD | COMP_HET | *PRKN* | Exon 2 deletion | Exon 4 deletion | | NCA, MLPA, WGS |
| 56 | PD | fEOPD | COMP HET | *PRKN* | G430D | N52Mfs*29 | | NCA, MLPA, WGS |
| | Unaffected | NA | HET | *PRKN* | G430D | - | | NCA, MLPA |
| 57 | PD | fEOPD | COMP_HET | *PRKN* | P113fs*51 | Exon 5 deletion | | NCA, MLPA, WGS |
| 58 | PD | sEOPD | COMP_HET | *PRKN* | Exons 3-4 deletion | Exon 8 deletion | *GBA1* T408M | NCA, MLPA, WGS |
| 59 | PD | fEOPD | COMP_HET | *PRKN* | Exon 3 deletion | Exons 5-7 deletion | | NCA, MLPA |
| 60 | PD | sEOPD | COMP_HET | *PRKN* | R275W | P113fs*51 | | NCA, MLPA, WGS |
| 61 | PD | fEOPD | COMP_HET | *PRKN* | R275W | c.1083+1G>T | | NCA, MLPA, WGS |
| | PD | fEOPD | COMP_HET | *PRKN* | R275W | c.1083+1G>T | | NCA, MLPA, WGS |
| 62 | PD | sEOPD | COMP_HET | *PRKN* | Exon 2 deletion | Exons 4-7 deletion | | NCA, MLPA, WGS |
| 63 | PD | sEOPD | COMP_HET | *PRKN* | Exon 3 duplication | Exon 7 duplication | | MLPA, WGS |
| 64 | PD | sEOPD | HOM | *PINK1* | Y258* | Y258* | | NCA, MLPA |
| 65 * | PD | fEOPD | HOM | *PINK1* | W90Lfs*12 | W90Lfs*12 | | NCA, MLPA, WGS |
| 66 | PD | sEOPD | HET | *SNCA* | Duplication | | | NCA, MLPA, WGS |
| 67 | PD | sEOPD | HET | *SNCA* | Duplication | | | NCA, MLPA |
| 68 | PD | fLOPD | HET | *VCP* | R159C | | | NCA, MLPA, WGS |
| 69 | PD | fEOPD | HET | *GCH1* | L224R | | *GBA1* R296Q | NCA, MLPA, WGS |

AAO = Age at onset, AAC = age at consent, NCA = NeuroChip array, MLPA = Multiplex ligation-dependent probe amplification, WGS = whole-genome sequencing, NGS = next-generation sequencing, AJ = Ashkenazi Jewish, EUR = European, SAS = South Asian, HET = heterozygous, HOM = homozygous, COMP_HET = compound heterozygous, NA = Not applicable. *Consanguineous family. ^Families found to be related by kinship analysis. †The frameshift deletion c.337_376del (P113fs*51) is located in exon 3 of the *PRKN* gene and disrupts the binding of MLPA exon 3 probe, causing a false positive exon 3 deletion in MLPA.

**Supplementary Table 2.** PD-associated variants with probes in NeuroChip genotyping array

**A. Variants detected at least once**

| Gene | Chr | Nucleotide change | Amino acid change | Clinvar classification | MDSGene classification | FP/total calls | FN/total calls |
|---|---|---|---|---|---|---|---|
| *PRKN* | 6 | c.1289G>A | p.Gly430Asp | Pathogenic | | 0/2 | 0/2 |
| *PRKN* | 6 | c.850G>C | p.Gly284Arg | Pathogenic | | 0/1 | 0/1 |
| *PRKN* | 6 | c.823C>T | p.Arg275Trp | Pathogenic | Definitely pathogenic | 0/8 | 0/8 |
| *PRKN* | 6 | c.633A>T | p.Lys211Asn | Pathogenic | Definitely pathogenic | 0/1 | 0/1 |
| *PRKN* | 6 | c.125G>C | p.Arg42Pro | Pathogenic | Definitely pathogenic | 0/1 | 0/1 |
| *PRKN* | 6 | c.1310C>T | p.Pro437Leu | Conflicting interpretations | Definitely pathogenic | 0/7 | 0/7 |
| *PRKN* | 6 | c.101_102del | p.Gln34fs* | Pathogenic | | 0/1 | 0/1 |
| *PRKN* | 6 | c.220_221dup | p.Trp74fs* | Pathogenic/Likely pathogenic | | 0/1 | 0/1 |
| *LRRK2* | 12 | c.4321C>T | p.Arg1441Cys | Pathogenic | Definitely pathogenic | 0/1 | 0/1 |
| *LRRK2* | 12 | c.6055G>A | p.Gly2019Ser | Pathogenic | Definitely pathogenic | 0/4 | 0/4 |
| *VCP* | 9 | c.475C>T | p.Arg159Cys | Pathogenic | | 0/1 | 0/1 |
| *GBA1* | 1 | c.1504C>T | p.Arg502Cys | Pathogenic | | 0/4 | 0/4 |
| *GBA1* | 1 | c.1342G>C | p.Asp448His | Pathogenic/Likely pathogenic | | 0/1 | 0/1 |
| *GBA1* | 1 | c.1246G>A | p.Gly416Ser | Pathogenic | | 0/1 | 0/1 |
| *GBA1* | 1 | c.1226A>G | p.Asn409Ser | Pathogenic/Likely pathogenic | | 0/3 | 0/3 |
| *GBA1* | 1 | c.1085C>T | p.Thr362Ile | Pathogenic | | 0/1 | 0/1 |
| *GBA1* | 1 | c.887G>A | p.Arg296Gln | Pathogenic | | 0/1 | 0/1 |
| *GBA1* | 1 | c.703T>C | p.Ser235Pro | Pathogenic | | **1/1** | 0/1 |
| *GBA1* | 1 | c.84dup | p.Leu29fs* | Pathogenic | | 0/1 | 0/1 |
| *GBA1* | 1 | c.475C>T | p.Arg159Trp | Pathogenic | | 0/1 | **1/1** |

**B. Variants not detected**

| Gene | Chr | c.DNA change | Amino acid change | Clinvar classification | MDSGene classification |
|---|---|---|---|---|---|
| *PRKN* | 6 | c.1359G>A | p.Trp453* | Likely pathogenic | Definitely pathogenic |
| *PRKN* | 6 | c.1292G>T | p.Cys431Phe | Pathogenic | |
| *PRKN* | 6 | c.931C>T | p.Gln311* | Pathogenic | Definitely pathogenic |
| *PRKN* | 6 | c.635G>A | p.Cys212Tyr | Pathogenic | Definitely pathogenic |
| *PRKN* | 6 | c.483A>T | p.Lys161Asn | Pathogenic | |
| *PRKN* | 6 | c.98G>A | p.Arg33Gln | Pathogenic | |
| *PRKN* | 6 | c.1334G>A | p.Trp445* | Pathogenic | |
| *PRKN* | 6 | c.1321T>C | p.Cys441Arg | Pathogenic | |
| *PRKN* | 6 | c.1252T>C | p.Cys418Arg | Likely pathogenic | |
| *PRKN* | 6 | c.1244C>A | p.Thr415Asn | Likely pathogenic | |
| *PRKN* | 6 | c.865T>G | p.Cys289Gly | | Definitely pathogenic |
| *PINK1* | 1 | c.502G>C | p.Ala168Pro | Pathogenic | Definitely pathogenic |
| *PINK1* | 1 | c.650C>A | p.Ala217Asp | Pathogenic | |
| *PINK1* | 1 | c.736C>T | p.Arg246* | Pathogenic | |
| *PINK1* | 1 | c.774C>A | p.Tyr258* | Pathogenic | |
| *PINK1* | 1 | c.813C>A | p.His271Gln | Pathogenic | Definitely pathogenic |
| *PINK1* | 1 | c.926G>A | p.Gly309Asp | Pathogenic | Definitely pathogenic |
| *PINK1* | 1 | c.938C>T | p.Thr313Met | Pathogenic | |
| *PINK1* | 1 | c.1040T>C | p.Leu347Pro | Pathogenic | Definitely pathogenic |
| *PINK1* | 1 | c.1311G>A | p.Trp437* | Pathogenic | Definitely pathogenic |
| *PINK1* | 1 | c.1366C>T | p.Gln456* | Pathogenic | Definitely pathogenic |
| *PINK1* | 1 | c.1474C>T | p.Arg492* | Pathogenic | |
| *PINK1* | 1 | c.1162T>C | p.Cys388Arg | Likely pathogenic | Definitely pathogenic |
| *PINK1* | 1 | c.377A>C | p.Gln126Pro | Likely pathogenic | |
| *PINK1* | 1 | c.509T>G | p.Val170Gly | | Definitely pathogenic |
| *PINK1* | 1 | c.718G>A | p.Glu240Lys | | Definitely pathogenic |
| *PINK1* | 1 | c.1226G>T | p.Gly409Val | | Definitely pathogenic |
| *PINK1* | 1 | c.1250A>G | p.Glu417Gly | | Definitely pathogenic |
| *PARK7* | 1 | c.29T>C | p.Leu10Pro | | Definitely pathogenic |
| *PARK7* | 1 | c.78G>A | p.Met26Ile | Pathogenic | Definitely pathogenic |
| *PARK7* | 1 | c.115G>T | p.Ala39Ser | Pathogenic | |
| *PARK7* | 1 | c.192G>C | p.Glu64Asp | Pathogenic | Definitely pathogenic |
| *PARK7* | 1 | c.497T>C | p.Leu166Pro | Pathogenic | Definitely pathogenic |
| *PARK7* | 1 | c.133C>T | p.Gln45* | Pathogenic | |
| *ATP13A2* | 1 | c.2629G>A | p.Gly877Arg | Pathogenic | |
| *ATP13A2* | 1 | c.2561T>G | p.Met854Arg | Pathogenic | |
| *ATP13A2* | 1 | c.1903C>T | p.Gln635* | Pathogenic | |
| *ATP13A2* | 1 | c.3176T>G | p.Leu1059Arg | Likely pathogenic | Definitely pathogenic |
| *ATP13A2* | 1 | c.2529+1G>A | | Likely pathogenic | |
| *ATP13A2* | 1 | c.477+2T>G | | Pathogenic/Likely pathogenic | |
| *ATP13A2* | 1 | c.2113C>T | p.Gln705Ter | Pathogenic | |
| *FBXO7* | 22 | c.65C>T | p.Thr22Met | Pathogenic | Definitely pathogenic |
| *FBXO7* | 22 | c.1492C>T | p.Arg498* | Pathogenic | Definitely pathogenic |
| *FBXO7* | 22 | c.961C>T | p.Arg321* | Likely pathogenic | |
| *FBXO7* | 22 | c.1033C>T | p.Arg345* | Likely pathogenic | |
| *SNCA* | 4 | c.157G>A | p.Ala53Thr | Pathogenic | Definitely pathogenic |
| *SNCA* | 4 | c.152G>A | p.Gly51Asp | Pathogenic | Definitely pathogenic |

| | | | | | |
|---|---|---|---|---|---|
| *SNCA* | 4 | c.136G>A | p.Glu46Lys | Pathogenic | |
| *SNCA* | 4 | c.88G>C | p.Ala30Pro | Pathogenic | |
| *LRRK2* | 12 | c.3364A>G | p.Ile1122Val | Pathogenic | |
| *LRRK2* | 12 | c.5096A>G | p.Tyr1699Cys | Pathogenic | Definitely pathogenic |
| *LRRK2* | 12 | c.6059T>C | p.Ile2020Thr | Pathogenic | Definitely pathogenic |
| *VPS35* | 16 | c.1858G>A | p.Asp620Asn | Pathogenic | Definitely pathogenic |
| *DCTN1* | 2 | c.221A>C | p.Gln74Pro | Pathogenic | |
| *DCTN1* | 2 | c.212G>A | p.Gly71Glu | Pathogenic | |
| *DCTN1* | 2 | c.211G>A | p.Gly71Arg | Pathogenic | Definitely pathogenic |
| *DCTN1* | 2 | c.200G>A | p.Gly67Asp | Pathogenic | |
| *DCTN1* | 2 | c.175G>A | p.Gly59Ser | Pathogenic | |
| *DCTN1* | 2 | c.156T>G | p.Phe52Leu | Pathogenic | |
| *DCTN1* | 2 | c.3823C>T | p.Arg1275Cys | Likely pathogenic | |
| *DCTN1* | 2 | c.175G>C | p.Gly59Arg | Pathogenic | |
| *VCP* | 9 | c.695C>A | p.Ala232Glu | Pathogenic | |
| *VCP* | 9 | c.572G>A | p.Arg191Gln | Pathogenic/Likely pathogenic | |
| *VCP* | 9 | c.476G>A | p.Arg159His | Pathogenic | |
| *VCP* | 9 | c.463C>T | p.Arg155Cys | Pathogenic | |
| *VCP* | 9 | c.283C>G | p.Arg95Gly | Pathogenic | |
| *VCP* | 9 | c.277C>T | p.Arg93Cys | Pathogenic | |
| *VCP* | 9 | c.472A>G | p.Met158Val | Pathogenic | |
| *VCP* | 9 | c.469G>C | p.Gly157Arg | Pathogenic | |
| *VCP* | 9 | c.284G>A | p.Arg95His | Likely pathogenic | |
| *VCP* | 9 | c.278G>A | p.Arg93His | Likely pathogenic | |
| *VCP* | 9 | c.553G>A | p.Glu185Lys | Likely pathogenic | |
| *VCP* | 9 | c.466G>A | p.Gly156Ser | Pathogenic | |
| *GBA1* | 1 | c.1549G>A | p.Gly517Ser | Pathogenic | |
| *GBA1* | 1 | c.1505G>A | p.Arg502His | Pathogenic/Likely pathogenic | |
| *GBA1* | 1 | c.1503C>G | p.Asn501Lys | Likely pathogenic | |
| *GBA1* | 1 | c.1448T>G | p.Leu483Arg | Pathogenic/Likely pathogenic | |
| *GBA1* | 1 | c.1297G>T | p.Val433Leu | Pathogenic/Likely pathogenic | |
| *GBA1* | 1 | c.1271T>C | p.Leu424Pro | Likely pathogenic | |
| *GBA1* | 1 | c.1228C>G | p.Leu410Val | Pathogenic | |
| *GBA1* | 1 | c.1208G>C | p.Ser403Thr | Likely pathogenic | |
| *GBA1* | 1 | c.1192C>T | p.Arg398* | Pathogenic | |
| *GBA1* | 1 | c.1174C>T | p.Arg392Trp | Pathogenic | |
| *GBA1* | 1 | c.1174C>G | p.Arg392Gly | Pathogenic | |
| *GBA1* | 1 | c.1171G>C | p.Val391Leu | Pathogenic/Likely pathogenic | |
| *GBA1* | 1 | c.1090G>A | p.Gly364Arg | Likely pathogenic | |
| *GBA1* | 1 | c.983C>T | p.Pro328Leu | Pathogenic | |
| *GBA1* | 1 | c.928A>G | p.Ser310Gly | Likely pathogenic | |
| *GBA1* | 1 | c.896T>C | p.Ile299Thr | Likely pathogenic | |
| *GBA1* | 1 | c.870C>A | p.Phe290Leu | Pathogenic | |
| *GBA1* | 1 | c.764T>A | p.Phe255Tyr | Pathogenic/Likely pathogenic | |
| *GBA1* | 1 | c.709A>G | p.Lys237Glu | Pathogenic | |
| *GBA1* | 1 | c.701G>A | p.Gly234Glu | Likely pathogenic | |
| *GBA1* | 1 | c.681T>G | p.Asn227Lys | Pathogenic | |
| *GBA1* | 1 | c.680A>G | p.Asn227Ser | Pathogenic/Likely pathogenic | |
| *GBA1* | 1 | c.625C>T | p.Arg209Cys | Pathogenic | |
| *GBA1* | 1 | c.509G>T | p.Arg170Leu | Likely pathogenic | |
| *GBA1* | 1 | c.497A>T | p.Asp166Val | Likely pathogenic | |
| *GBA1* | 1 | c.415G>C | p.Ala139Pro | Likely pathogenic | |
| *GBA1* | 1 | c.254G>A | p.Gly85Glu | Pathogenic | |
| *GBA1* | 1 | c.160G>T | p.Val54Leu | Pathogenic | |
| *GBA1* | 1 | c.914C>T | p.Pro305Leu | Likely pathogenic | |
| *GBA1* | 1 | c.894C>A | p.Phe298Leu | Likely pathogenic | |
| *GBA1* | 1 | c.1249T>G | p.Trp417Gly | Pathogenic/Likely pathogenic | |
| *GBA1* | 1 | c.661C>A | p.Pro221Thr | Pathogenic/Likely pathogenic | |
| *GBA1* | 1 | c.314T>C | p.Leu105Pro | Likely pathogenic | |
| *GBA1* | 1 | c.1300C>T | p.Arg434Cys | Likely pathogenic | |
| *GBA1* | 1 | c.1162G>A | p.Glu388Lys | Likely pathogenic | |
| *GBA1* | 1 | c.721G>A | p.Gly241Arg | Pathogenic/Likely pathogenic | |

Pathogenic variants in genes associated with monogenic PD, classified by ClinVar as pathogenic/likely pathogenic and/or MDSGene as definitively pathogenic. *GBA1* variants classified by ClinVar as pathogenic for Gaucher's disease or risk factors for PD. The proportion of false positive (FP) and false negative (FN) genotype calls were calculated for the 20 probes targeting mutations detected in the 537 individuals who underwent both NeuroChip array genotyping and sequencing. There were 40/42 (95.2%) matched genotypes and 90% (18/20) of probes had a sensitivity and specificity of 100%.

**Supplementary Table 3.** Clinical features of index cases with pathogenic variants in PD-related genes

| | *SNCA* | *GCH1* | *ATXN2* | *VCP* | *PLA2G6* | *PNPLA6* | *SPG7* | *PINK1* |
|---|---|---|---|---|---|---|---|---|
| Index cases (N) | 2 | 1 | 3 | 1 | 1 | 1 | 1 | 2 |
| Age at Onset (mean ± sd) * | 40.0 (5.7) | 31-35 | 56.3 (3.79) | 56-60 | 31-35 | 26-30 | 21-25 | 36.5 (12.0) |
| Motor Features (mean ± sd) | | | | | | | | |
| MDS-UPDRS Part III | 49 | 31 | 45 (NA) | 38 | NA | 23 | NA | 16 |
| Motor Severity Score | 4.1 | 3.4 | 3.7 (NA) | 4.7 | NA | 2.9 | NA | 0.6 |
| Motor Subtype (%) | | | | | | | | |
| Tremor-dominant | 0 | 100 | 0 | 0 | 0 | 0 | NA | NA |
| PIGD-dominant | 100 | 0 | 100 | 100 | 0 | 100 | NA | NA |
| Intermediate | 0 | 0 | 0 | 0 | 0 | 0 | NA | NA |
| Hoehn and Yahr stage (%) | | | | | | | | |
| 0-1.5 | 0 | 0 | 100 | 0 | NA | 0 | NA | 0 |
| 2 or 2.5 | 0 | 100 | 0 | 100 | NA | 100 | NA | 100 |
| 3+ | 100 | 0 | 0 | 0 | NA | 0 | NA | 0 |
| Motor Complications (%) | | | | | | | | |
| Dyskinesias | 100 | 0 | 0 | 0 | NA | 100 | NA | 100 |
| Motor fluctuations | 100 | 100 | 100 | 100 | NA | 0 | NA | 0 |
| Off dystonia | 100 | 100 | 0 | 0 | NA | 0 | NA | 0 |
| Motor Aspects of Daily Living (mean ± sd) | 28 | 20 | 15.3 (11.1) | 17 | 16 | 20 | NA | NA |
| Autonomic Dysfunction (%) | | | | | | | | |
| Orthostatic Hypotension | 100 | 100 | 66.7 | 100 | 100 | 100 | 100 | 100 |
| Constipation | 100 | 100 | 100 | 0 | 100 | 0 | 100 | 100 |
| Urinary Dysfunction | 100 | 100 | 100 | 0 | 100 | 0 | 100 | 100 |
| REM Sleep Behaviour Disorder (%) | 100 | 100 | 66.7 | 0 | 100 | 100 | 0 | 100 |
| Neuropsychiatric Symptoms (%) | | | | | | | | |
| Apathy | 100 | 100 | 50 | 0 | NA | 0 | 0 | 0 |
| Depression | 100 | 100 | 0 | 0 | NA | 0 | 0 | 0 |
| Anxiety | 100 | 100 | 0 | 0 | NA | 0 | 0 | 0 |
| Dopamine Dysregulation Syndrome | 100 | 100 | 0 | 0 | NA | 0 | 0 | 0 |
| Hallucinations | 100 | 100 | 0 | 100 | NA | 0 | 0 | 0 |
| MoCA score (mean ± sd) | 27 | 26 | 18.5 (0.7) | 30 | 28 | 26 | 25 | 23 |

MDS-UPDRS items were used to define the following clinical features: dyskinesias (items 4.1+4.2 >0); motor fluctuations (items 4.3+4.4+4.5 >0); off-dystonia (item 4.6 >0); orthostatic hypotension (item 1.12 >0 ); constipation (item 1.11 >0); urinary dysfunction (item 1.10 >0); apathy (item 1.5 >0); depression (item 1.3 >1); anxiety (item 1.4 >1), impulse control disorder (item 1.6 >0); hallucinations (item 1.2 >0 and <4). Motor severity scores are MDS-UPDRD part III scores divided by disease duration in years. Motor subtypes were defined according to Stebbins et al., 2013. Motor aspects of daily living scores are the sum of MDS-UPDRS part II items. REM sleep behaviour disorder was defined as RBDSQ score >5. * When N = 1, Age at Onset is presented as a 5-year age bracket.

**Supplementary Table 4.** Variant details of families carrying a single heterozygous pathogenic variant in *PINK1* and *PRKN*

| Family ID | Diagnosis | Group | Gene | Variant 1 | Variant 2 | Risk variants | Investigations |
|---|---|---|---|---|---|---|---|
| | PD | fEOPD | *PINK1* | Exon 5 deletion | | *GBA* N409S | NCA, MLPA, WGS |
| 1 | Unaffected | NA | - | - | | *GBA* N409S | NCA, MLPA |
| | Unaffected | NA | *PINK1* | Exon 5 deletion | | - | NCA, MLPA |
| 2 | PD | fLOPD | *PINK1* | c.1124-1G>A | | | NCA, MLPA, WGS |
| 3 | PD | fLOPD | *PRKN* | K211N | | *GBA1* E365K | NCA, MLPA, WGS |
| 4 | PD | fEOPD | *PRKN* | P437L | | | NCA, MLPA, WGS |
| 5 | PD | fEOPD | *PRKN* | P437L | - | | NCA, MLPA, WGS |
| | PD | fLOPD | *PRKN* | P437L | P132Tfs*9 | | NCA, MPLA, WGS |
| 6 | PD | fLOPD | *PRKN* | P437L | | | NCA, MLPA, WGS |
| 7 | PD | fLOPD | *PRKN* | P437L | | | NCA, MLPA, WGS |
| 8 | PD | fLOPD | *PRKN* | P437L | | | NCA, MLPA, WGS |
| 9 | PD | fLOPD | *PRKN* | R275W | | *GBA1* T408M | NCA, MLPA, WGS |
| 10 | PD | fLOPD | *PRKN* | Exons 1-2 duplication | | | NCA, MLPA, WGS |
| 11 | PD | sEOPD | *PRKN* | Exon 1 deletion | | *GBA1* L483P | NCA, MLPA, WGS |
| 12 | PD | fLOPD | *PRKN* | Exon 2 deletion | | | NCA, MLPA, WGS |
| 13 | PD | sEOPD | *PRKN* | Exon 3 deletion | | | NCA, MLPA, WGS |
| 14 | PD | fLOPD | *PRKN* | Exons 3-6 duplication | | | NCA, MLPA, WGS |

AAO = Age at onset, AAC = age at consent, NCA = NeuroChip array, MLPA = Multiplex ligation-dependent probe amplification, WGS = whole-genome sequencing, EUR = European CAS = Central Asian, NA = Not applicable.
Only index cases with completed MLPA and WGS were included.

**Supplementary Table 5.** Unique variants identified in index cases

| MOI | Gene | Variant | Allele frequency (N alleles) sEOPD | fEOPD | fLOPD | Total |
|---|---|---|---|---|---|---|
| *Autosomal dominant* | | | | | | |
| | *LRRK2* | G2019S | 1.2 (5) | 0.9 (2) | 2.6 (21) | 1.9 (28) |
| | *LRRK2* | R1441C | 0 | 0 | 0.1 (1) | 0.1 (1) |
| | *LRRK2* | N1437S | 0 | 0 | 0.1 (1) | 0.1 (1) |
| | *ATXN2* | CAG repeat | 0 | 0 | 0.4 (3) | 0.2 (3) |
| | *SNCA* | Duplication | 0.5 (2) | 0 | 0 | 0.1 (2) |
| | *VCP* | R159C | 0 | 0 | 0.1 (1) | 0.1 (1) |
| | *GCH1* | L224R | 0 | 0.4 (1) | 0 | 0.1 (1) |
| | *RAB32* | S71R | 0 | 0 | 0.1 (1) | 0.1 (1) |
| *Autosomal recessive* | | | | | | |
| | *PARK2* | R275W | 2 (8) | 2.2 (5) | 0.1 (1) | 1 (14) |
| | *PARK2* | P437L | 0.2 (1) | 1.3 (3) | 0.8 (6) | 0.7 (10) |
| | *PARK2* | P113fs | 1 (4) | 0.4 (1) | 0 | 0.3 (5) |
| | *PARK2* | G430D | 0.2 (1) | 0.9 (2) | 0 | 0.2 (3) |
| | *PARK2* | **C166Hfs*18** | 0.2 (1) | 0 | 0 | 0.1 (1) |
| | *PARK2* | C166Y | 0 | 0.4 (1) | 0 | 0.1 (1) |
| | *PARK2* | C253Y | 0 | 0.4 (1) | 0 | 0.1 (1) |
| | *PARK2* | G284R | 0.2 (1) | 0 | 0 | 0.1 (1) |
| | *PARK2* | K211N | 0 | 0 | 0.1 (1) | 0.1 (1) |
| | *PARK2* | N52fs | 0.2 (1) | 0.4 (1) | 0 | 0.1 (2) |
| | *PARK2* | Q34Rfs | 0.5 (2) | 0 | 0 | 0.1 (2) |
| | *PARK2* | R42P | 0 | 0.4 (1) | 0 | 0.1 (1) |
| | *PARK2* | T240M | 0.2 (1) | 0 | 0 | 0.1 (1) |
| | *PARK2* | W74Cfs*8 | 0.2 (1) | 0 | 0 | 0.1 (1) |
| | *PARK2* | **c.1083+1G>T** | 0 | 0.4 (1) | 0 | 0.1 (1) |
| | *PARK2* | Exon 2 deletion | 1.2 (5) | 0 | 0.1 (1) | 0.4 (6) |
| | *PARK2* | Exon 3 deletion | 0.2 (1) | 0.9 (2) | 0 | 0.2 (3) |
| | *PARK2* | Exon 4 deletion | 0.5 (2) | 0.4 (1) | 0 | 0.2 (3) |
| | *PARK2* | Exon 8 deletion | 0.5 (2) | 0.4 (1) | 0 | 0.2 (3) |
| | *PARK2* | Exon 2 duplication | 0 | 0 | 0.2 (2) | 0.1 (2) |
| | *PARK2* | Exon 3 duplication | 0.2 (1) | 0.4 (1) | 0 | 0.1 (2) |
| | *PARK2* | Exons 3-4 deletion | 0.2 (1) | 0.4 (1) | 0 | 0.1 (2) |
| | *PARK2* | Exons 3-6 duplication | 0 | 0.4 (1) | 0.1 (1) | 0.1 (2) |
| | *PARK2* | Exon 5 deletion | 0 | 0.9 (2) | 0 | 0.1 (2) |
| | *PARK2* | Exon 1 deletion | 0.2 (1) | 0 | 0 | 0.1 (1) |
| | *PARK2* | Exons 1-2 duplication | 0 | 0 | 0.1 (1) | 0.1 (1) |
| | *PARK2* | Exons 1-3 deletion | 0 | 0.4 (1) | 0 | 0.1 (1) |
| | *PARK2* | Exons 4-7 deletion | 0.2 (1) | 0 | 0 | 0.1 (1) |
| | *PARK2* | Exons 5-7 deletion | 0 | 0.4 (1) | 0 | 0.1 (1) |
| | *PARK2* | Exons 7 duplication | 0.2 (1) | 0 | 0 | 0.1 (1) |
| | *PINK1* | W90Lfs*12 | 0 | 0.9 (2) | 0 | 0.1 (2) |
| | *PINK1* | Y258* | 0.5 (2) | 0 | 0 | 0.1 (2) |
| | *PINK1* | **c.1124-1G>A** | 0 | 0 | 0.1 (1) | 0.1 (1) |
| | *PINK1* | Exon 5 deletion | 0 | 0.9 (2) | 0 | 0.1 (2) |
| | *PNPLA6* | P1297S | 0.5 (2) | 0 | 0 | 0.1 (2) |
| | *PLA2G6* | L354P | 0.2 (1) | 0 | 0 | 0.1 (1) |
| | *PLA2G6* | T319M | 0.2 (1) | 0 | 0 | 0.1 (1) |
| | *SPG7* | A510V | 0 | 0.4 (1) | 0 | 0.1 (1) |
| | *SPG7* | N288* | 0 | 0.4 (1) | 0 | 0.1 (1) |
| *PD risk* | | | | | | |
| | *GBA1* | E365K | 1.5 (6) | 1.8 (4) | 1.6 (13) | 1.6 (23) |
| | *GBA1* | T408M | 1.5 (6) | 1.8 (4) | 1 (8) | 1.3 (18) |
| | *GBA1* | L483P | 1.5 (6) | 0 | 0.8 (6) | 0.8 (12) |
| | *GBA1* | N409S | 0.2 (1) | 0.9 (2) | 0.2 (2) | 0.3 (5) |
| | *GBA1* | A163fs | 0 | 0 | 0.1 (1) | 0.1 (1) |
| | *GBA1* | D448H | 0 | 0 | 0.1 (1) | 0.1 (1) |
| | *GBA1* | **G152A** | 0.2 (1) | 0 | 0 | 0.1 (1) |
| | *GBA1* | G152E | 0.2 (1) | 0 | 0 | 0.1 (1) |
| | *GBA1* | G416S | 0 | 0.4 (1) | 0 | 0.1 (1) |
| | *GBA1* | IVS2DS+1G-A | 0 | 0.4 (1) | 0 | 0.1 (1) |
| | *GBA1* | L29Afs*18 | 0 | 0 | 0.1 (1) | 0.1 (1) |
| | *GBA1* | M162T | 0 | 0 | 0.1 (1) | 0.1 (1) |
| | *GBA1* | P138Lfs*62 | 0.5 (2) | 0 | 0 | 0.1 (2) |
| | *GBA1* | **P430H** | 0 | 0 | 0.1 (1) | 0.1 (1) |
| | *GBA1* | **P454A** | 0.2 (1) | 0 | 0 | 0.1 (1) |
| | *GBA1* | R159Q | 0 | 0 | 0.1 (1) | 0.1 (1) |
| | *GBA1* | R159W | 0 | 0.4 (1) | 0 | 0.1 (1) |
| | *GBA1* | R296Q | 0 | 0.4 (1) | 0 | 0.1 (1) |
| | *GBA1* | R398* | 0 | 0 | 0.1 (1) | 0.1 (1) |
| | *GBA1* | R502C | 0.2 (1) | 0 | 0.4 (3) | 0.3 (4) |

**Supplementary Table 5.** Unique variants identified in index cases

| MOI | Gene | Variant | Allele frequency (N alleles) | | | |
|---|---|---|---|---|---|---|
| | | | **sEOPD** | **fEOPD** | **fLOPD** | **Total** |
| *PD risk* | *GBA1* | T362I | 0 | 0.4 (1) | 0 | 0.1 (1) |
| | *GBA1* | **Y244Tfs*10** | 0 | 0 | 0.1 (1) | 0.1 (1) |
| | *GBA1* | **Y352*** | 0 | 0 | 0.1 (1) | 0.1 (1) |
| | *GBA1* | c.1263del | 0 | 0.4 (1) | 0 | 0.1 (1) |
| | *GBA1* | **V333Rfs*4** | 0 | 0.4 (1) | 0 | 0.1 (1) |

sEOPD = sporadic early-onset PD, fEOPD = familial early-onset PD, fLOPD = familial late-onset PD, MOI = mode of inheritance.
Bold indicates novel variants. An additional novel *PRKN* variant (P132Tfs*9) was exclusively identified in an affected relative and is not listed here.

**Supplementary Table 6.** Frequency of pathogenic and risk factor variants identified in index cases

| | sEOPD<br>N = 205 | fEOPD<br>N = 113 | fLOPD<br>N = 400 | Total<br>N = 718 |
|---|---|---|---|---|
| **PD-causing genes (%)** | **12.2** | **14.2** | **7** | **9.6** |
| *LRRK2* | 2.4 | 1.8 | 5.8 | 4.2 |
| *SNCA* | 1 | 0 | 0 | 0.3 |
| *VCP* | 0 | 0 | 0.2 | 0.1 |
| *ATXN2* | 0 | 0 | 0.8 | 0.4 |
| *GCH1* | 0 | 0.9 | 0 | 0.1 |
| *RAB32* | 0 | 0 | 0.2 | 0.1 |
| *PRKN* (biallelic) | 7.3 | 9.7 | 0 | 3.6 |
| *PINK1* (biallelic) | 0.5 | 0.9 | 0 | 0.3 |
| *PNPLA6* (biallelic) | 0.5 | 0 | 0 | 0.1 |
| *PLA2G6* (biallelic) | 0.5 | 0 | 0 | 0.1 |
| *SPG7* (biallelic) | 0 | 0.9 | 0 | 0.1 |
| **Single recessive variants (%) ‡** | **1.3** | **3.3** | **3** | **1.9** |
| *PRKN* (monoallelic) | 1.3 | 2.2 | 2.7 | 1.7 |
| *PINK1* (monoallelic) | 0 | 1.1 | 0.3 | 0.3 |
| ***GBA1* risk variants (%) †** | **10.2** | **13.3** | **9.2** | **10.2** |
| PD risk variants | 4.9 | 7.1 | 4.5 | 5 |
| Mild GD-causing mutations | 0.5 | 1.8 | 0.5 | 0.7 |
| Severe GD-causing mutations | 2.9 | 3.5 | 3.2 | 3.2 |
| Unknown severity | 2 | 0.9 | 1 | 1.3 |

‡Excludes single recessive pathogenic variants in cases not fully investigated with MLPA and WGS (n=9). †Excludes *GBA1* mutations that coexist with pathogenic mutations in *LRRK2* (*n*=1), *PRKN* biallelic (*n*=2), *PRKN* monoallelic (n=3), *PINK1* monoallelic (*n*=1) and *GCH1* (*n*=1). sEOPD = sporadic early-onset PD, fEOPD = familial early-onset PD, fLOPD = familial late-onset PD, GD = Gaucher disease.

**Supplementary Table 7.** Demographic characteristics of index cases by gene

| | *SNCA* | *VCP* | *ATXN2* | *RAB32* | *LRRK2* | *PNPLA6* | *PINK1* | *SPG7* | *PLA2G6* | *PRKN* | *GCH1* | *GBA1* † | Mutation-negative ‡ | P-value |
|---|---|---|---|---|---|---|---|---|---|---|---|---|---|---|
| Index cases (N) | 2 | 1 | 3 | 1 | 30 | 1 | 2 | 1 | 1 | 26 | 1 | 70 | 431 | |
| Sex (N Female:Male) | 1:1 | 0:1 | 3:0 | 1:0 | 15:15 | 0:1 | 2:0 | 0:1 | 1:0 | 10:16 | 0:1 | 28:42 | 185:246 | 0.269 |
| Age at Diagnosis (mean ± sd) ***** | 44.5 (0.7) d | 61-65 | 63.3 (4.2) d | 55-60 | 59.5 (13.7) d | 31-35 | 44.5 (6.4) d | 41-45 | 36-40 | 36.4 (8.0) **c** | 36-40 | 53.9 (12.5) d | 55.8 (14.0) | **8.52E-09** |
| Age at Assessment (mean ± sd) ***** | 48.5 (0.7) d | 66-70 | 76.0 (5.2) d | 55-60 | 64.5 (11.6) d | 36-40 | 54.5 (0.7) d | 46-50 | 41-45 | 50.0 (12.3) **a** | 41-45 | 58.6 (12.2) d | 60.6 (13.2) | **2.11E-04** |
| Disease duration at assessment (Years, mean ± sd) | 8.5 (4.9) d | 8 | 19.7 (7.5) d | 1 | 6.8 (4.9) d | 8 | 18 (11.3) d | 25 | 9 | 21.6 (14.0) **c** | 9 | 9.5 (9.8) d | 8.4 (8.3) | **1.69E-06** |
| Genetically Determined Ancestry (%) | | | | | | | | | | | | | | 0.367 |
| African | 0 | 0 | 0 | 0 | 0 | 0 | 0 | 0 | 0 | 0 | 0 | 1.4 | 0.7 | |
| American | 0 | 0 | 0 | 0 | 0 | 0 | 0 | 0 | 0 | 0 | 0 | 0 | 0.7 | |
| Ashkenazi Jewish | 0 | 0 | 0 | 0 | 10 | 100 | 0 | 0 | 0 | 0 | 0 | 1.4 | 1.2 | |
| Central Asian | 0 | 0 | 0 | 0 | 0 | 0 | 0 | 0 | 0 | 0 | 0 | 0 | 0.5 | |
| East Asian | 0 | 0 | 0 | 0 | 0 | 0 | 0 | 0 | 0 | 0 | 0 | 0 | 0.2 | |
| European | 100 | 100 | 100 | 100 | 90 | 0 | 50 | 100 | 100 | 100 | 100 | 91.4 | 91.4 | |
| Finnish | 0 | 0 | 0 | 0 | 0 | 0 | 0 | 0 | 0 | 0 | 0 | 1.4 | 0 | |
| Middle Eastern | 0 | 0 | 0 | 0 | 0 | 0 | 0 | 0 | 0 | 0 | 0 | 0 | 0.5 | |
| South Asian | 0 | 0 | 0 | 0 | 0 | 0 | 50 | 0 | 0 | 0 | 0 | 2.9 | 4.4 | |
| Complex Admixture | 0 | 0 | 0 | 0 | 0 | 0 | 0 | 0 | 0 | | 0 | 0 | 0.2 | |
| Unknown | 0 | 0 | 0 | 0 | 0 | 0 | 0 | 0 | 0 | 0 | 0 | 1.4 | 0.5 | |
| Family history (%) | | | | | | | | | | | | | | 0.001** |
| No family history | 100 | 0 | 0 | 0 | 16.7 | 100 | 50 | 0 | 100 | 57.7 | 0 | 28.6 | 28.8 | |
| One affected relative | 0 | 0 | 33.3 | 0 | 30 | 0 | 50 | 100 | 0 | 30.8 | 100 | 40.0 | 48.0 | |
| Two affected relatives | 0 | 100 | 33.3 | 100 | 26.7 | 0 | 0 | 0 | 0 | 7.7 | 0 | 20.0 | 16.0 | |
| Three or more affected relatives | 0 | 0 | 33.3 | 0 | 26.7 | 0 | 0 | 0 | 0 | 3.8 | 0 | 11.4 | 7.2 | |
| Self-reported parental consanguinity (%) | 0 | 0 | 0 | 0 | 3.4 | 0 | 100 | 0 | 0 | 4.3 | 0 | 0 | 1.3 | 0.015 ** |

†Excludes cases not investigated with WGS (*n*=3) and *GBA1* variants that coexist with pathogenic mutations in *LRRK2* (*n*=1), *PRKN* biallelic (*n*=2), *PRKN* monoallelic (*n*=3), *PINK1* monoallelic (*n*=1) and *GCH1* (*n*=1).

‡ Excludes mutation-negative cases not investigated with both WGS and MLPA (*n*=122), carriers of *GBA1* variants and of monoallelic pathogenic *PRKN* and *PINK1* mutations.

* When N = 1, Age at Diagnosis and Age at Assessment are presented as a 5-year age bracket.

Continuous variables were tested with Kruskal-Wallis test with FDR-adjusted P-values for multiple comparisons. Categorical variables were tested with Fisher's exact test.

Only genes with N≥2 were tested. Significance levels show comparison with mutation-negative group only: a, P<0.05. b, P<0.001. c, P<0.0001. d, not significant. **No significant P-values after FDR-adjusted pairwise comparisons across genes.

**Supplementary Table 8.** Clinical features of *PRKN* single mutation carriers index cases

| | | ***PRKN* monoallelic vs mutation-negative ‡** | | ***PRKN* monoallelic vs *PRKN* biallelic** | |
|---|---|---|---|---|---|
| | ***PRKN* monoallelic †** | **Beta (95% CI)** | **P-value** | **Beta (95% CI)** | **P-value** |
| Age at Onset (mean ± sd) | 52.2 (15.5) | -2.9 (-10.4, 4.6) | 0.449 | 10.4 (1.1, 19.6) | **0.028** |
| Age at Diagnosis (mean ± sd) | 54.8 (15.2) | -2.4 (-10.2, 5.3) | 0.535 | 12.0 (2.5, 21.6) | **0.013** |
| Age at Assessment (mean ± sd) | 57.2 (14.9) | -2.9 (-10.4, 4.6) | 0.449 | 10.4 (1.1, 19.7) | **0.028** |
| Disease duration at assessment (Years, mean ± sd) | 4.9 (2.6) | -3.3 (-8.3, 1.6) | 0.187 | -16.6 (-22.5, -10.7) | **6.21E-08** |
| MDS-UPDRS Part III (mean ± sd) | 19.6 (8.8) | -3.7 (-13.2, 5.8) | 0.447 | 1.0 (-11.6, 13.5) | 0.881 |
| Motor Severity Score (mean ± sd) | 6.1 (8.4) | 0.02 (-4.0, 4.0) | 0.992 | 4.4 (-0.6, 9.4) | 0.083 |
| Motor Aspects of Daily Living (mean ± sd) | 9.8 (9.1) | 2.0 (-6.9, 3) | 0.431 | 7.1 (1.0, 13.1) | **0.022** |
| Motor Complications (%) | | | | | |
| Dyskinesias | 27.3 | 0.36 (-1.2, 1.66) | 0.610 | 1.4 (-0.6, 3.4) | 0.155 |
| Motor fluctuations | 27.3 | -0.4 (-2.0, 0.9) | 0.548 | 1.3 (-0.8, 3.3) | 0.230 |
| Off dystonia | 9.1 | -0.9 (-3.8, 0.84) | 0.411 | 0.3 (-2.9, 2.6) | 0.822 |
| Autonomic Dysfunction (%) | | | | | |
| Orthostatic Hypotension | 45.5 | -0.10 (-1.4, 1.1) | 0.869 | 0.8 (-0.7, 2.3) | 0.298 |
| Constipation | 27.3 | -0.8 (-2.4, 0.5) | 0.239 | 0.0 (-1.8, 1.6) | 0.995 |
| Urinary Dysfunction | 54.5 | -0.19 (-1.4, 1.1) | 0.756 | 0.6 (-0.9, 2.2) | 0.412 |
| REM Sleep Behaviour Disorder (%) | 45.5 | 0.4 (-0.8, 1.6) | 0.483 | 0.8 (-0.6, 2.3) | 0.259 |
| Neuropsychiatric Symptoms (%) | | | | | |
| Apathy | 27.3 | 0.0 (-1.6, 1.2) | 0.951 | 0.8 (-1.1, 2.6) | 0.400 |
| Depression | 18.2 | 0.3 (-1.6, 1.7) | 0.712 | 1.2 (-1.1, 3.6) | 0.282 |
| Anxiety | 27.3 | 0.4 (-1.1, 1.7) | 0.528 | 0.4 (-1.4, 2.2) | 0.620 |
| Dopamine Dysregulation Syndrome | 9.1 | -0.3 (-3.3, 1.4) | 0.752 | 0.3 (-2.9, 2.6) | 0.833 |
| Hallucinations | 18.2 | 0.4 (-1.5, 1.8) | 0.606 | 1.8 (-0.6, 4.2) | 0.125 |
| Total MoCA score (mean ± sd) | 27.0 (4.4) | 0.27 (-1.6, 2.2) | 0.778 | 0.2 (-2.2, 2.7) | 0.856 |

† Excludes cases without both MLPA and WGS. ‡ Excludes mutation-negative cases not investigated with both WGS and MLPA (*n*=122), carriers of *GBA1* variants and of monoallelic pathogenic *PRKN* and *PINK1* mutations. Linear, logistic or multinomial regression as appropriate, after adjustment for sex, age and disease duration (except age at onset, which was adjusted only for sex and disease duration, and motor severity, which was adjusted only for sex and age). Significance level set at <0.05.

**Supplementary Table 9.** Variant details of families carrying a pathogenic or risk variant in *GBA1*

| Family ID | Diagnosis | Group | Gene | Variant | Additional variants | Investigations |
|---|---|---|---|---|---|---|
| 1 | PD | fLOPD | *GBA1* | D448H | | NCA, MLPA, WGS |
| 2 | PD | sEOPD | *GBA1* | G152E | E365K | NCA, MLPA, WGS |
| 3 | PD | sEOPD | *GBA1* | G152A | | NCA, MLPA, WGS |
| 4 | PD | fEOPD | *GBA1* | G416S | | NCA, MLPA, WGS |
| 5 | PD | fLOPD | *GBA1* | N409S | | NCA, MLPA |
| 6 | PD | fEOPD | *GBA1* | N409S | *PINK1* exon 5 deletion | NCA, MLPA, WGS |
| | Unaffected | NA | *GBA1* | N409S | | NCA, MLPA |
| | Unaffected | NA | - | - | *PINK1* exon 5 deletion | NCA, MLPA |
| 7 | PD | fLOPD | *GBA1* | N409S | | NCA, MLPA, WGS |
| 8 | PD | fEOPD | *GBA1* | N409S | | NCA, MLPA, WGS |
| | Unaffected | NA | *GBA1* | N409S | | NCA |
| 9 | PD | sEOPD | *GBA1* | N409S | | NCA, MLPA |
| 10 | PD | sEOPD | *GBA1* | P138Lfs*62 | | WGS |
| 11 | PD | sEOPD | *GBA1* | P138Lfs*62 | | NCA, MLPA, WGS |
| 12 | PD | fLOPD | *GBA1* | Y244Tfs*10 | | NCA, MLPA, WGS |
| 13 | PD | fEOPD | *GBA1* | V333Rfs*4 | | NCA, MLPA, WGS |
| 14 | PD | fLOPD | *GBA1* | A163fs | | NCA, MLPA, WGS |
| 15 | PD | fEOPD | *GBA1* | c.1263del | | NCA, MLPA, WGS |
| | PD | fLOPD | *GBA1* | c.1263del | | NCA, MLPA, WGS |
| 16 | PD | fEOPD | *GBA1* | IVS2DS+1G-A | | NCA, MLPA, WGS |
| 17 | PD | fLOPD | *GBA1* | P430H | | NCA, WGS |
| 18 | PD | fLOPD | *GBA1* | R159Q | | NCA, MLPA, WGS |
| 19 | PD | fEOPD | *GBA1* | R159W | | NCA, MLPA, WGS |
| 20 | PD | fEOPD | *GBA1* | R296Q | *GCH1* K224R | NCA, MLPA, WGS |
| | Unaffected | NA | *GBA1* | R296Q | | NCA |
| | Unaffected | NA | *GBA1* | R296Q | | NCA |
| 21 | PD | fLOPD | *GBA1* | R398* | | NCA, MLPA |
| 22 | PD | fLOPD | *GBA1* | Y352* | | NCA, MLPA, WGS |
| 23 | PD | fLOPD | *GBA1* | R502C | | NCA, MLPA, WGS |
| 24 | PD | fLOPD | *GBA1* | R502C | | NCA, MLPA, WGS |
| 25 | PD | fLOPD | *GBA1* | R502C | | NCA, MLPA, WGS |
| 26 | PD | sEOPD | *GBA1* | R502C | | NCA, MLPA, WGS |
| 27 | PD | fEOPD | *GBA1* | T362I | | NCA, MLPA, WGS |
| 28 | PD | fLOPD | *GBA1* | L29Afs*18 | *LRRK2* G2019S | NCA, MLPA, WGS |
| 29 | PD | fLOPD | GBA1 | E365K | | NCA, MLPA, WGS |
| 30 | PD | fLOPD | GBA1 | E365K | | NCA, MLPA, WGS |
| 31 | PD | fLOPD | GBA1 | L483P | | NCA, MLPA, WGS |
| 32 | PD | fLOPD | *GBA1* | T408M | | NCA, MLPA, WGS |
| 33 | PD | fLOPD | *GBA1* | T408M | | NCA, MLPA, WGS |
| 34 | PD | fLOPD | *GBA1* | E365K | | NCA, MLPA, WGS |
| 35 | PD | fLOPD | *GBA1* | T408M | | NCA, MLPA, WGS |
| 36 | PD | sEOPD | *GBA1* | L483P | | NCA, MLPA, WGS |
| 37 | PD | fLOPD | *GBA1* | T408M | E365K | NCA, MLPA, WGS |
| | PD | fLOPD | *GBA1* | T408M | E365K | NCA, MLPA, WGS |
| 38 | PD | fEOPD | *GBA1* | T408M | | NCA, MLPA, WGS |
| | Unaffected | NA | - | - | | NCA, MLPA |
| 39 | PD | fLOPD | *GBA1* | E365K | | NCA, MLPA, WGS |
| 40 | PD | sEOPD | *GBA1* | T408M | | NCA, MLPA, WGS |
| 41 | PD | sEOPD | *GBA1* | L483P | | NCA, MLPA, WGS |
| | Unaffected | NA | - | - | | NCA |
| 42 | PD | sEOPD | *GBA1* | T408M | | NCA, MLPA, WGS |
| | Unaffected | NA | - | - | | NCA |
| | Unaffected | NA | - | - | | NCA |
| 43 | PD | sEOPD | *GBA1* | L483P | | NCA, MLPA, WGS |
| 44 | PD | fEOPD | *GBA1* | T408M | | NCA, MLPA, WGS |
| 45 | PD | fLOPD | *GBA1* | E365K | | NCA, MLPA, WGS |
| 46 | PD | fEOPD | *GBA1* | T408M | | NCA, WGS |
| 47 | PD | sEOPD | *GBA1* | L483P | | NCA, MLPA, WGS |
| 48 | PD | fEOPD | *GBA1* | E365K | | NCA, MLPA, WGS |
| 49 | PD | sEOPD | *GBA1* | E365K | | NCA, MLPA, WGS |
| 50 | PD | fLOPD | *GBA1* | E365K | | NCA, MLPA, WGS |
| 51 | PD | fLOPD | *GBA1* | M162T | | NCA, MLPA, WGS |
| 52 | PD | sEOPD | *GBA1* | E365K | | NCA, MLPA, WGS |
| 53 | PD | sEOPD | *GBA1* | E365K | | NCA, MLPA, WGS |
| 54 | PD | fLOPD | *GBA1* | L483P | | NCA, MLPA, WGS |
| 55 | PD | sEOPD | *GBA1* | T408M | | NCA, MLPA, WGS |
| 56 | PD | sEOPD | *GBA1* | E365K | | NCA, MLPA, WGS |
| 57 | PD | fLOPD | *GBA1* | T408M | | NCA, MLPA, WGS |
| 58 | PD | fLOPD | *GBA1* | L483P | | NCA, MLPA, WGS |

**Supplementary Table 9.** Variant details of families carrying a pathogenic or risk variant in *GBA1*

| Family ID | Diagnosis | Group | Gene | Variant | Additional variants | Investigations |
|---|---|---|---|---|---|---|
| 59 | PD | fLOPD | *GBA1* | E365K | | NCA, MLPA, WGS |
| 60 | PD | fLOPD | *GBA1* | L483P | | NCA, MLPA, WGS |
| 61 | PD | fLOPD | *GBA1* | E365K | | NCA, MLPA, WGS |
| 62 | PD | fEOPD | *GBA1* | E365K | | NCA, MLPA, WGS |
| 63 | PD | fLOPD | *GBA1* | L483P | | NCA, MLPA, WGS |
| 64 | PD | fEOPD | *GBA1* | T408M | | NCA, MLPA, WGS |
| 65 | PD | fLOPD | *GBA1* | L483P | | NCA, MLPA, WGS |
| | Unaffected | NA | - | - | | NCA, MLPA |
| 66 | PD | sEOPD | *GBA1* | T408M | | NCA, MLPA, WGS |
| 67 | PD | fEOPD | *GBA1* | E365K | | NCA, MLPA, WGS |
| | Unaffected | NA | - | - | | NCA, MLPA |
| 68 | PD | sEOPD | *GBA1* | P454A | | NCA, MLPA, WGS |
| | Unaffected | NA | - | - | | NCA |
| 69 | PD | fLOPD | *GBA1* | T408M | | NCA, MLPA, WGS |
| 70 | PD | fEOPD | *GBA1* | E365K | | NCA, MLPA, WGS |
| | Unaffected | NA | - | - | | NCA, MLPA |
| 71 | PD | fLOPD | *GBA1* | E365K | | NCA, MLPA, WGS |
| 72 | PD | fLOPD | *GBA1* | E365K | | NCA, MLPA, WGS |
| 73 | PD | fLOPD | *GBA1* | T408M | | NCA, MLPA, WGS |
| | Unaffected | NA | - | - | | NCA, MLPA |
| | Unaffected | NA | - | - | | NCA, MLPA |
| | Unaffected | NA | - | - | | NCA, MLPA |
| 74 | PD | fLOPD | *GBA1* | E365K | | NCA, MLPA, WGS |
| 75 | PD | sEOPD | *GBA1* | L483P | | MLPA, WGS |
| 76 | PD | sEOPD | *GBA1* | E365K | | MLPA, WGS |
| 77 | PD | fLOPD | *GBA1* | T408M | *PRKN* R275W | NCA, MLPA, WGS |
| 78 | PD | fLOPD | *GBA1* | E365K | *PRKN* K211N | NCA, MLPA, WGS |
| 79 | PD | sEOPD | *GBA1* | L483P | *PRKN* exon 1 deletion | NCA, MLPA, WGS |
| 80 | PD | sEOPD | *GBA1* | T408M | *PRKN* G248R | NCA, WGS |
| 81 | PD | sEOPD | *GBA1* | T408M | *PRKN* exons 3-4 deletion | NCA, MLPA, WGS |

AAO = Age at onset, AAC = age at consent, NCA = NeuroChip array, MLPA = Multiplex ligation-dependent probe amplification, WGS = whole-genome sequencing, AJ = Ashkenazi Jewish, EUR = European, SAS = South Asian, FIN = Finnish, AFR = African, NA = Not applicable.

**Supplementary Table 10.** Clinical features of *GBA1* mutation-carriers vs mutation-negative by variant severity in index cases

| | Risk N = 34 | Mild (N = 3) | Severe (N = 22) | Mutation-negative ‡ N = 431 | P-value | | |
|---|---|---|---|---|---|---|---|
| | | | | | Risk | Mild | Severe |
| Age at Onset (mean ± sd) | 51.6 (16.2) | 44.3 (16.0) | 47.4 (13.9) | 52.3 (15.0) | 0.884 | 0.625 | 0.337 |
| Motor Features (mean ± sd) | | | | | | | |
| MDS-UPDRS Part III | 25.2 (13.2) | 39.0 (11.3) | 27.5 (17.9) | 26.5 (17.4) | 0.696 | 0.484 | 0.672 |
| Motor Severity Score | 6.6 (6.0) | 4.7 (4.4) | 6.1 (6.7) | 6.3 (6.7) | 0.788 | 0.726 | 0.814 |
| Motor Subtype (%) | | | | | | | |
| Tremor-dominant | 28.6 | 0 | 30.8 | 39.1 | | | |
| PIGD-dominant | 61.9 | 100 | 61.5 | 52.1 | 0.451 | NA | 0.524 |
| Intermediate | 9.5 | 0 | 7.7 | 8.8 | 0.659 | NA | 0.901 |
| Hoehn and Yahr stage (%) | | | | | | | |
| 0-1.5 | 45.8 | 0 | 7.7 | 38.2 | | | |
| 2 or 2.5 | 29.2 | 0 | 61.5 | 37.8 | 0.346 | NA | 0.075 |
| 3+ | 25 | 100 | 30.8 | 24 | 0.583 | NA | 0.149 |
| Motor Complications (%) | | | | | | | |
| Dyskinesias | 26.1 | 50 | 23.1 | 26 | 0.729 | 0.808 | 0.415 |
| Motor fluctuations | 45.5 | 100 | 25 | 43.4 | 0.920 | 0.985 | **0.048** |
| Off dystonia | 19 | 100 | 14.3 | 24 | 0.484 | 0.985 | 0.159 |
| Motor Aspects of Daily Living (mean ± sd) | 13.2 (8.9) | 19.3 (14.0) | 12.7 (7.6) | 13.5 (9.2) | 0.714 | 0.459 | 0.248 |
| Autonomic Dysfunction (%) | | | | | | | |
| Orthostatic Hypotension | 35.5 | 66.7 | 47.6 | 48.6 | 0.154 | 0.708 | 0.907 |
| Constipation | 68.8 | 66.7 | 57.1 | 49.1 | **0.034** | 0.644 | 0.385 |
| Urinary Dysfunction | 64.5 | 66.7 | 61.9 | 64.2 | 0.932 | 0.995 | 0.813 |
| REM Sleep Behaviour Disorder (%) | 50 | 66.7 | 52.4 | 39.9 | | | |
| Neuropsychiatric Symptoms (%) | | | | | | | |
| Apathy | 12.5 | 50 | 40 | 30.7 | 0.055 | 0.727 | 0.543 |
| Depression | 0 | 50 | 18.8 | 14.8 | 0.984 | 0.318 | 0.918 |
| Anxiety | 8.3 | 50 | 18.8 | 20.8 | 0.141 | 0.446 | 0.625 |
| Dopamine Dysregulation Syndrome | 4.3 | 50 | 31.2 | 13.2 | 0.205 | 0.266 | 0.125 |
| Hallucinations | 12.5 | 50 | 31.2 | 15.5 | 0.603 | 0.395 | 0.242 |
| MoCA score (mean ± sd) | 26.3 (2.8) | 24.5 (3.5) | 24.7 (3.1) | 26.3 (3.4) | 0.847 | 0.477 | **0.039** |

Excludes *GBA1* variants of unknown severity (*n*=9), *GBA1* mutations that coexist with pathogenic mutations in other PD-related genes (*n*=8), individuals not investigated with WGS (*n*=3) and individuals who are compound heterozygous for *GBA1* variants (*n*=2). ‡ Excludes mutation-negative cases not investigated with both WGS and MLPA (*n*=122), carriers of *GBA1* variants and of monoallelic pathogenic *PRKN* and *PINK1* mutations. Linear, logistic or multinomial regression were used as appropriate, after adjustment for sex, age and disease duration (except age at onset, which was adjusted only for sex and disease duration, and motor severity, which was adjusted only for sex and age). Significance level set at <0.05. MDS-UPDRS items were used to define the following clinical features: dyskinesias (items 4.1+4.2 >0); motor fluctuations (items 4.3+4.4+4.5 >0); off-dystonia (item 4.6 >0); orthostatic hypotension (item 1.12 >0 ); constipation (item 1.11 >0); urinary dysfunction (item 1.10 >0); apathy (item 1.5 >0); depression (item 1.3 >1); anxiety (item 1.4 >1), impulse control disorder (item 1.6 >0); hallucinations (item 1.2 >0 and <4). Motor severity scores are MDS-UPDRD part III scores divided by disease duration in years. Motor subtypes were defined according to Stebbins et al., 2013. Motor aspects of daily living scores are the sum of MDS-UPDRS part II items. REM sleep behaviour disorder was defined as RBDSQ score >5. NA=not applicable.

**Supplementary Table 11.** GP2 gene panel

| Gene | Ensembl gene ID | MANE Select |
|---|---|---|
| AFG3L2 | ENSG00000141385 | NM_006796.3 |
| ATP13A2 | ENSG00000159363 | NM_022089.4 |
| ATP1A3 | ENSG00000105409 | NM_152296.5 |
| ATP6AP2 | ENSG00000182220 | NM_005765.3 |
| ATP7B | ENSG00000123191 | NM_000053.4 |
| C19orf12 | ENSG00000131943 | NM_031448.6 |
| CHCHD10 | ENSG00000250479 | NM_213720.3 |
| CHCHD2 | ENSG00000106153 | NM_016139.4 |
| COASY | ENSG00000068120 | NM_025233.7 |
| CP | ENSG00000047457 | NM_000096.4 |
| CSF1R | ENSG00000182578 | NM_001288705.3 |
| DCAF17 | ENSG00000115827 | NM_025000.4 |
| DCTN1 | ENSG00000204843 | NM_004082.5 |
| DNAJC12 | ENSG00000108176 | NM_021800.3 |
| DNAJC6 | ENSG00000116675 | NM_001256864.2 |
| EPM2A | ENSG00000112425 | NM_005670.4 |
| FA2H | ENSG00000103089 | NM_024306.5 |
| FBXO7 | ENSG00000100225 | NM_012179.4 |
| FTL | ENSG00000087086 | NM_000146.4 |
| GBA1 | ENSG00000177628 | NM_000157.4 |
| GCH1 | ENSG00000131979 | NM_000161.3 |
| GLB1 | ENSG00000170266 | NM_000404.4 |
| GRN | ENSG00000030582 | NM_002087.4 |
| JAM2 | ENSG00000154721 | NM_021219.4 |
| LRRK2 | ENSG00000188906 | NM_198578.4 |
| LYST | ENSG00000143669 | NM_000081.4 |
| MAPT | ENSG00000186868 | NM_001377265.1 |
| MYORG | ENSG00000164976 | NM_020702.5 |
| NPC1 | ENSG00000141458 | NM_000271.5 |
| NPC2 | ENSG00000119655 | NM_006432.5 |
| NR4A2 | ENSG00000153234 | NM_006186.4 |
| OPA1 | ENSG00000198836 | NM_130837.3 |
| OPA3 | ENSG00000125741 | NM_025136.4 |
| PANK2 | ENSG00000125779 | NM_001386393.1 |
| PARK7 | ENSG00000116288 | NM_007262.5 |
| PDE8B | ENSG00000113231 | NM_003719.5 |
| PDGFB | ENSG00000100311 | NM_002608.4 |
| PDGFRB | ENSG00000113721 | NM_002609.4 |
| PINK1 | ENSG00000158828 | NM_032409.3 |
| PLA2G6 | ENSG00000184381 | NM_003560.4 |
| POLG | ENSG00000140521 | NM_002693.3 |
| PRKN | ENSG00000185345 | NM_004562.3 |
| PRKRA | ENSG00000180228 | NM_003690.5 |
| PTRHD1 | ENSG00000184924 | NM_001013663.2 |
| PTS | ENSG00000150787 | NM_000317.3 |
| QDPR | ENSG00000151552 | NM_000320.3 |
| RAB39B | ENSG00000155961 | NM_171998.4 |
| SCP2 | ENSG00000116171 | NM_002979.5 |
| SLC20A2 | ENSG00000168575 | NM_001257180.2 |
| SLC25A4 | ENSG00000151729 | NM_001151.4 |
| SLC25A46 | ENSG00000164209 | NM_138773.4 |
| SLC30A10 | ENSG00000196660 | NM_018713.3 |
| SLC39A14 | ENSG00000104635 | NM_001128431.4 |
| SLC6A3 | ENSG00000142319 | NM_001044.5 |
| SMPD1 | ENSG00000166311 | NM_000543.5 |
| SNCA | ENSG00000145335 | NM_000345.4 |
| SPG11 | ENSG00000104133 | NM_025137.4 |
| SPG7 | ENSG00000197912 | NM_003119.4 |
| SPR | ENSG00000116096 | NM_003124.5 |

**Supplementary Table 11.** GP2 gene panel

| Gene | Ensembl gene ID | MANE Select |
|---|---|---|
| SQSTM1 | ENSG00000161011 | NM_003900.5 |
| SYNJ1 | ENSG00000159082 | NM_203446.3 |
| TAF1 | ENSG00000147133 | NM_004606.5 |
| TBK1 | ENSG00000183735 | NM_013254.4 |
| TH | ENSG00000180176 | NM_000360.4 |
| TUBB4A | ENSG00000104833 | NM_006087.4 |
| TWNK | ENSG00000107815 | NM_021830.5 |
| VAC14 | ENSG00000103043 | NM_018052.5 |
| VCP | ENSG00000165280 | NM_007126.5 |
| VPS13A | ENSG00000197969 | NM_033305.3 |
| VPS13C | ENSG00000129003 | NM_020821.3 |
| VPS35 | ENSG00000069329 | NM_018206.6 |
| WDR45 | ENSG00000196998 | NM_001029896.2 |
| XPR1 | ENSG00000143324 | NM_004736.4 |
| ZFYVE26 | ENSG00000072121 | NM_015346.4 |

**Supplementary Table 12.** Primer Sequences

| | Forward (5' – 3') | Reverse (5' – 3') |
|---|---|---|
| ***ATXN2*** | 6-FAM-TTTCGGCGGCTCCTTGGTCTC | AGCCGCGGGCGGCGGCTGCTGCTGCTGCTG |
| ***ATXN3*** | 6-FAM-AGTCCAGTGACTACTTTGATTCG | GTCCTGATAGGTCCCCCTGCTGCTGCTGCTG |

### PFP Study Group

**Lead Investigators:** Prof Huw R. Morris (UCL Queen Square Institute of Neurology), Dr Paul R. Jarman (National Hospital for Neurology and Neurosurgery), Prof Henry Houlden (UCL Queen Square Institute of Neurology), Prof Nicholas W. Wood (UCL Queen Square Institute of Neurology), Dr Raquel Real (UCL Queen Square Institute of Neurology)

**Co-Investigators:** Manuela Tan (UCL Queen Square Institute of Neurology), Russel Tilney (UCL Queen Square Institute of Neurology)

**Principal investigators for sites:** Huw R. Morris (Royal Free London NHS Trust and University College London Hospitals NHS Foundation Trust), Elizabeth Wakeman (Isle of Wight NHS Trust), Tabish Saifee (London North West Healthcare NHS Trust), Sam Arianayagam (Hampshire Hospitals NHS Foundation Trust), Saifuddin Shaik (Lancashire Teaching Hospitals NHS Trust), Sophie Molloy (Imperial College Healthcare NHS Trust and London North West Healthcare NHS Trust), Ralph Gregory (University Hospitals Dorset NHS Foundation Trust), Mirdhu Wickremaratchi (Western Sussex Hospitals NHS Foundation Trust), Rosaria Buccoliero (Harrogate and District NHS Foundation Trust), Oliver Bandmann (Sheffield Teaching Hospital NHS Foundation Trust), Dominic Paviour (St George's University Hospitals NHS Foundation Trust), Diran Padiachy (Salisbury NHS Foundation Trust), Anjum Misbahuddin (Barking, Havering and Redbridge University Hospitals NHS Trust), Jeremy Cosgrove (Leeds Teaching Hospitals NHS Trust), Sunku Guptha (North West Anglia NHS Foundation Trust), Ray Chaudhuri (King's College Hospital NHS Foundation Trust), Yen Tai (The Hillingdon Hospitals NHS Foundation Trust), Sukaina Asad (University Hospitals of Leicester NHS Trust), Ayano Funaki (Central London Community Healthcare NHS Trust ), Marek Kunc (Airedale NHS Foundation Trust), Charlotte Brierley (West Suffolk NHS Foundation Trust), Ray Sheridan (Royal Devon & Exeter NHS Foundation Trust), Rena Truscott (Livewell Southwest), Suzanne Dean (Royal Cornwall Hospitals NHS Trust), Carinna Vickers (Somerset NHS Foundation Trust), Rani Sophia (Yeovil District Hospital NHS Foundation Trust), Sion Jones (Betsi Cadwaladr University Health Board), Erica Capps (The Shrewsbury and Telford Hospital NHS Trust), Neil Archibald (South Tees Hospitals NHS Foundation Trust), Louise Wiblin (South Tees Hospitals NHS Foundation Trust), Sean J Slaght (Portsmouth Hospitals NHS Trust), Edward Jones (York Teaching Hospital NHS Foundation Trust), Colin Barnes (Solent NHS Trust), Dominick D'Costa (The Royal Wolverhampton NHS Trust), Carl Mann (Midlands Partnership NHS Foundation Trust), Uma Nath (South Tyneside and Sunderland NHS Foundation Trust), Anette Schrag (Bedfordshire Hospitals NHS Foundation Trust), Sarah Williams (St Helens and Knowsley Teaching Hospitals NHS Trust), Gillian Webster (North Cumbria Integrated Care NHS Foundation Trust), Sigurlaug Sveinbjornsdottir (Mid and South Essex NHS Foundation Trust), Lucy Strens (University Hospitals Coventry & Warwickshire), Annette Hand (Northumbria Healthcare NHS Foundation Trust), Richard Walker (Northumbria Healthcare NHS Foundation Trust), Rosemary Crouch (Birmingham Community Healthcare NHS Trust), Jason Raw (Northern Care Alliance NHS Foundation Trust), Stephanie Tuck (Norfolk Community Health & Care NHS Trust), Khaled Amar (University Hospital Dorset NHS Foundation Trust), Emma Wales (Wye Valley NHS Trust), Irene Gentilini (Cambridge University Hospitals NHS

Foundation Trust), Aileen Nacorda (Cambridge University Hospitals NHS Foundation Trust), Louise Hartley (Barts Health NHS Trust)

**Clinical research assistants and support staff for sites:** Simona Jasaityte (Royal Free London NHS Trust and University College London Hospitals NHS Foundation Trust, Research Assistant), Miriam Pollard (Royal Free London NHS Trust and University College London Hospitals NHS Foundation Trust, Research Assistant), Megan Hodgson (University College London Hospitals NHS Foundation Trust, Research Assistant), Clodagh Towns (Royal Free London NHS Trust and University College London Hospitals NHS Foundation Trust, Research Assistant), Matilda Fenn (Royal Free London NHS Trust and University College London Hospitals NHS Foundation Trust, Research Assistant), Sarah Cable (Royal Free London NHS Trust and University College London Hospitals NHS Foundation Trust, Research Assistant), Kaylee Gauntlett (Isle of Wight NHS Trust), Deborah Dellafera (Hampshire Hospitals NHS Foundation Trust, Senior Research Nurse), Vicki Fleming (Lancashire Teaching Hospitals NHS Trust, Research Nurse), Judith Dube (University Hospitals Dorset NHS Foundation Trust, Research Nurse), Sarah House (Western Sussex Hospitals NHS Foundation Trust, Clinical Research Practitioner), Caroline Bennett (Harrogate and District NHS Foundation Trust), Helen Casbolt (Sheffield Teaching Hospital NHS Foundation Trust, Clinical Trials Assistant), James Lee (Salisbury NHS Foundation Trust), Laura Parker-Wall (Barking, Havering and Redbridge University Hospitals NHS Trust, Research Nurse), Prisca Mpofu (Leeds Teaching Hospitals NHS Trust, Research Nurse), Dhaval Trivedi (King's College Hospital NHS Foundation Trust), Sofiya Portukhay (The Hillingdon Hospitals NHS Foundation Trust, Clinical Trials Coordinator), Rehanah Roopun (University Hospitals of Leicester NHS Trust, Research Nurse), Lisa Armstrong (Airedale NHS Foundation Trust, Clinical Trials Coordinator), Clare O'Reilly (Royal Devon & Exeter NHS Foundation Trust, Senior Research Nurse), Jonathan Gilby (Livewell Southwest, Senior Research Practitioner), Nishi Singh (Somerset NHS Foundation Trust, Research Nurse), Alison Lewis (Yeovil District Hospital NHS Foundation Trust, research nurse), Julia Roberts (Betsi Cadwaladr University Health Board), Claire Watkins (Betsi Cadwaladr University Health Board), Hannah Gibson (The Shrewsbury and Telford Hospital NHS Trust, Assistant Research Practitioner), Jo Stickley (The Shrewsbury and Telford Hospital NHS Trust, Research Nurse), Catherine Edwards (Portsmouth Hospitals NHS Trust, PD Nurse Specialist), Kerry Elliott (York Teaching Hospital NHS Foundation Trust, Research Nurse), Emma Searle (Solent NHS Trust, Research Nurse), Blessing Okoko (The Royal Wolverhampton NHS Trust, Research Nurse), Susan Wilson (South Tyneside and Sunderland NHS Foundation Trust, Research Nurse), Yvynne Croucher (Bedfordshire Hospitals NHS Foundation Trust), Sharon Dealing (St Helens and Knowsley Teaching Hospitals NHS Trust, Research Nurse), Hannah Martin (University Hospitals Coventry & Warwickshire, Advanced Nurse Practitioner), Claire Williams (Birmingham Community Healthcare NHS Trust, Research Nurse), Lucy Booth (Birmingham Community Healthcare NHS Trust, Research Nurse), Steph Howard (Norfolk Community Health & Care NHS Trust), Rochelle Hernandez (University Hospital Dorset NHS Foundation Trust, Research Nurse), Sue Anderson (Wye Valley NHS Trust, Clinical Trials Nurse), Nosheen Khalid (Barts Health NHS Trust, Clinical Trial Practitioner)

### GP2 Consortium

| Affiliation | Contributor |
| --- | --- |
| Centre Hospitalo-Universitaire Dr Benbadis Constantine, Constantine, Algeria | Yasser Mecheri |
| Sanatorio de la Trinidad Mitre- INEBA, Buenos Aires, Argentina | Emilia M Gatto |
| Hospital JM Ramos Mejia, Buenos Aires, Argentina | Marcelo Kauffman |
| Centro de Educación Médica e Investigaciones Clínicas Norberto Quirno, Buenos Aires, Argentina | Federico Capparelli |
| Somnus Neurology Clinic, Yerevan, Armenia | Samson Khachatryan, Zaruhi Tavadyan, Mariam Isayan |
| Neuroscience Research Australia, Sydney, New South Wales, Australia | Claire E Shepherd, Simon Rowe, Dennis Yeow Carolyn Sue |
| ANZAC Research Institute, Concord, New South Wales, Australia | Julie Hunter |
| Garvan Institute of Medical Research, Darlinghurst, New South Wales, Australia | Kishore Kumar |
| Concord Hospital, Concord, New South Wales, Australia | Melina Ellis, Kishore Kumar |
| QIMR Berghofer Medical Research Institute, Herston, Queensland, Australia | Miguel E. Rentería, Victor Flores Ocampo |
| Murdoch University, Perth, Western Australia, Australia | Sulev Koks |
| Medical University Vienna Austria, Vienna, Austria | Alexander Zimprich |
| Istanbul Klinik, Baku, Azerbaijan | Kanan Jafarov |
| University of Antwerp, Antwerp, Belgium | David Crosiers |
| Universidade Federal do Rio Grande do Sul, Porto Alegre, Brazil | Artur F. Schumacher-Schuh, Paula Saffie Awad, |
| Hospital de Clínicas de Porto Alegre, Porto Alegre, Brazil | Artur F. Schumacher-Schuh |
| Federal University of Health Sciences of Porto Alegre, Porto Alegre, Brazil | Carlos Rieder |
| University of São Paulo, São Paulo, Brazil | Vitor Tumas |
| Universidade Federal de Minas Gerais , Belo Horizonte, Brazil | Sarah Camargos, Lucas Faria Costa |
| Montreal Neurological Institute, Montreal, Quebec, Canada | Edward A. Fon |
| Institut universitaire de gériatrie de Montréal , Montreal, Quebec, Canada | Oury Monchi |
| McGill University, Montreal, Quebec, Canada | Ted Fon, Ziv Gan-Or, Konstantin Senkevich |
| Aligning Science Across Parkinson's, Vancouver, British Columbia, Canada | Robert Thibault, Devin Sharp |
| University of Toronto, Toronto, Ontario, Canada | Anthony Lang |
| Universidad de Chile, Santiago, Chile | Benjamin Pizarro Galleguillos, Patricio Olguin, Maria Leonor Bustamante |
| Fundación Diagnosis, Santiago, Chile | Marcelo Miranda |
| CETRAM, Santiago, Chile | Pedro Chana |
| Hospital Regional de Concepción, Concepcion, Chile | Elias Fernandez |
| Central South University, Changsha, China | Beisha Tang, Zhenhua Liu |
| West China Hospital Sichuan University, Chengdu, China | Huifang Shang |
| Xiangya Hospital, Changsha, China | Zhenhua Liu, Jifeng Guo |
| Capital Medical University, Beijing, China | Piu Chan |
| Zhejiang University, Hangzhou, China | Wei Luo |
| Universidad Nacional de Colombia, Bogotá, Colombia | Gonzalo Arboleda |

| Affiliation | Contributor |
|---|---|
| Fundación Valle del Lili, Santiago De Cali, Colombia | Jorge Orozco |
| University of Antioquia, Medellin, Colombia | Marlene Jimenez del Rio |
| University of Costa Rica, San Jose, Costa Rica | Alvaro Hernandez |
| Aarhus University, Aarhus, Denmark | Per Borghammer |
| The American University in Cairo, Cairo, Egypt | Mohamed Salama |
| Beni-Suef University, Beni Suef, Egypt | Walaa A. Kamel |
| Addis Ababa University, Addis Ababa, Ethiopia | Yared Z. Zewde |
| Paris Brain Institute, Paris, France | Alexis Brice |
| Sorbonne Université, Paris, France | Jean-Christophe Corvol, Mari Vidailhet |
| Salpêtriere Hospital (AP-HP), Paris, France | Mari Vidailhet |
| Tbilisi State Medical University, Tbilisi, Georgia | Mariam Kekenadze |
| S. Khechinashvili University Hospital, Tbilisi, Georgia | Irine Khatiashvili |
| University of Lübeck, Lübeck, Germany | Ana Westenberger, Christine Klein, Eva-Juliane Vollstedt, Harutyun Madoev, Joanne Trinh, Johanna Junker, Katja Lohmann, Theresa Luth, Inke König, Lara M. Lange |
| University Medical Center Göttingen, Göttingen, Germany | Brit Mollenhauer |
| Department of Neurology, University Hospital, LMU Munich, Munich, Germany | Franziska Hopfner, Günter Höglinger |
| University Medical Center Schleswig-Holstein, Lübeck, Germany | Lara M. Lange, Daniela Berg |
| University of Tubingen, Tübingen, Germany | Manu Sharma, Thomas Gasser, Wenhua Sun |
| University of Mainz, Mainz, Germany | Sergiu Groppa |
| The German Center for Neurodegenerative Diseases, Tübingen, Germany | Zih-Hua Fang, Anastasia Illarionova |
| Charité - Universitätsmedizin Berlin, Berlin, Germany | Karl Heilbron |
| Technical University of Munich, Munich, Germany | Bernhard Haslinger |
| University of Ghana Medical School, Accra, Ghana | Albert Akpalu |
| University of Thessaly, Volos, Greece | Georgia Xiromerisiou, Georgios Hadjigorgiou, Efthymios Dadiotis |
| Aristotle University of Thessaloniki, Thessaloniki, Greece | Ioannis Dagklis |
| Ionian University, Corfu, Greece | Ioannis Tarnanas |
| Biomedical research Foundation of the Academy of Athens, Athens, Greece | Leonidas Stefanis |
| Diagnostic and Therapeutic Centre HYGEIA Hospital, Marousi, Greece | Maria Stamelou |
| Hospital San Felipe, Tegucigalpa, Honduras | Alex Medina |
| Queen Elizabeth Hospital , Kowloon, Hong Kong | Germaine Hiu-Fai Chan, Nelson Yuk-Fai Cheung |
| The Hong Kong University of Science and Technology, Kowloon, Hong Kong | Nancy Ip, Phillip Chan , Xiaopu Zhou |
| Aster Medcity, Kochi, India | Asha Kishore |
| Sree Chitra Tirunal Institute for Medical Sciences and Technology, Thiruvananthapuram, India | Divya KP |
| National Institute of Mental Health & Neurosciences, Bengaluru, India | Pramod Pal |
| Manipal Hospital, Delhi, India | Prashanth Lingappa Kukkle |

| Affiliation | Contributor |
|---|---|
| All India Institute of Medical Sciences, Delhi, India | Roopa Rajan |
| Nizam's Institute Of Medical Sciences, Hyderabad, India | Rupam Borgohain |
| Shahid Beheshti University of Medical Science, Tehran, Iran | Mehri Salari |
| Tel Aviv Sourasky Medical Center, Tel Aviv-Yafo, Israel | Tamara Shiner, Avner Thaler |
| Magna Græcia University of Catanzaro, Catanzaro, Italy | Andrea Quattrone, Monica Gagliardi |
| University of Pavia, Pavia, Italy | Enza Maria Valente |
| National Research Council, Cosenza, Italy | Grazia Annesi |
| University of Perugia, Perugia, Italy | Lucilla Parnetti |
| University of Rome Tor Vergata, Rome, Italy | Tommaso Schirinzi |
| IRCCS Mondino Foundation, Pavia, Italy | Caterina Galandra |
| University of Naples Federico II, Naples, Italy | Anna De Rosa |
| Juntendo University, Tokyo, Japan | Manabu Funayama, Nobutaka Hattori |
| Jikei University School of Medicine, Tokyo, Japan | Tomotaka Shiraishi |
| Institute of Neurology and Neurorehabilitation, Almaty, Kazakhstan | Altynay Karimova, Gulnaz Kaishibayeva |
| West Kazakhstan Marat Ospanov State Medical University, Aktobe, Kazakhstan | Aigerim Utegenova, Aigul Yemagambetova |
| Medline Medical Center, Astana, Kazakhstan | Vadim Akhmetzhanov |
| National Center for Neurosurgery, Astana, Kazakhstan | Seitzhan Aidarov |
| Astana Medical University, Astana, Kazakhstan | Tautanova Raushan, Dinara Alzhanova |
| City multi-field hospital No. 1, Astana, Kazakhstan | Bagyzhan Syzdykova Syzdykova |
| International University of Postgraduate Education, Almaty, Kazakhstan | Zhanybek Myrzayev |
| South Kazakhstan Medical Academy, Shymkent, Kazakhstan | Saltanat Abdraimova |
| Kyrgyz State Medical Academy, Bishkek, Kyrgyzstan | Cholpon Shambetova |
| University of Luxembourg, Esch-sur-Alzette, Luxembourg | Rejko Krüger, Patrick May |
| University of Malaya, Kuala Lumpur, Malaysia | Ai Huey Tan, Azlina Ahmad-Annuar, Shen-Yang Lim, Yi Wen Tay, Azalea Tenerife Pajo |
| Universiti Kebangsaan Malaysia, Selangor, Malaysia | Mohamed Ibrahim Norlinah |
| UKM Medical Molecular Biology Institute, Kuala Lumpur, Malaysia | Nor Azian Abdul Murad |
| Universiti Kebangsaan Malaysia Medical Centre, Kuala Lumpur, Malaysia | Shahrul Azmin |
| International Islamic University, Kuala Lumpur, Malaysia | Wael Mohamed |
| Tecnologico de Monterrey, Monterrey, Mexico | Daniel Martinez-Ramirez |
| Instituto Nacional de Neurologia y Neurocirugia, Mexico City, Mexico | Mayela Rodriguez-Violante, Nancy Monroy Jaramillo |
| Universidad Nacional Autónoma de México, Santiago de Querétaro, Mexico | Paula Reyes-Pérez, Alejandra Medina Rivera |
| Mongolian National University of Medical Sciences, Ulaanbaatar, Mongolia | Bayasgalan Tserensodnom |
| Tribhuvan University, Kirtipur, Nepal | Rajeev Ojha |
| Vanderbilt University Medical Center, Amsterdam, Netherlands | Wilma Van De Berg |
| Radboud University, Nijmegen, Netherlands | Bas Bleom, Bart Van de Warrenburg |

| Affiliation | Contributor |
|---|---|
| Brain Research and Innovation Center, Amsterdam, Netherlands | Lisette Charbonnier |
| University of Otago, Dunedin, New Zealand | Tim J. Anderson, Toni L. Pitcher |
| University of Lagos, Lagos, Nigeria | Arinola Sanyaolu, Njideka Okubadejo, Oluwadamilola Ojo |
| University of Calabar Teaching Hospital, Calabar, Nigeria | SIMON IZUCHUKWU OZOMMA |
| University of Ilorin, Ilorin, Nigeria | Kolawole Wahab |
| Oslo University Hospital, Oslo, Norway | Lasse Pihlstrøm, Manuela Tan, Ingeborg Haugesag Lie |
| Stavanger University Hospital, Stavanger, Norway | Jodi Maple-Grødem |
| University of Science and Technology Bannu, Bannu, Pakistan | Shoaib Ur-Rehman |
| Universidad Cientifica del Sur, Lima, Peru | Mario Cornejo-Olivas |
| Metropolitan Medical Center, Manila, Philippines | Maria Leila Doquenia, Raymond Rosales |
| University of Puerto Rico, San Juan, Puerto Rico | Angel Vinuela |
| Research Center of Neurology, Moscow, Russia | Elena Iakovenko |
| Ufa Federal Research Center, Ufa, Russia | Anna Gareeva |
| Ufa Scientific Center, Ufa, Russia | Gulnara Akhmadeeva |
| Bashkir State Medical University, Ufa, Russia | Irina Gilyazova |
| King Faisal Specialist Hospital and Research Center, Riyadh, Saudi Arabia | Bashayer Al Mubarak |
| King Abdullah International Medical Research Center, Jeddah, Saudi Arabia | Muhammad Umair |
| National Neuroscience Institute , Singapore, Singapore | Eng-King Tan |
| Nanyang Technological University, Singapore, Singapore | Jia Nee Foo, Elaine Chew |
| Ljubljana University Medical Centre, Ljubljana, Slovenia | Vesna van Midden |
| University of KwaZulu-Natal, Durban, South Africa | Ferzana Amod |
| University of Stellenbosch, Stellenbosch, South Africa | Jonathan Carr, Soraya Bardien, Kathryn Step |
| University of the Western Cape, Bellville, South Africa | Nikita Pillay |
| Seoul National University Hospital, Seoul, South Korea | Beomseok Jeon |
| Yongin Severance Hospital, Seoul, South Korea | Yun Joong Kim |
| Seoul National University, Seoul, South Korea | Jung Hwan Shin, Joowon Jang |
| Hospital Universitario Burgos, Burgos, Spain | Esther Cubo |
| University Hospital Mutua Terrassa, Barcelona, Spain | Ignacio Alvarez |
| Institut de Recerca Sant Joan de Deu, Barcelona, Spain | Janet Hoenicka |
| Research Institute Germans Trias i Pujol, Barcelona, Spain | Katrin Beyer |
| Instituto de Biomedicina de Sevilla, Seville, Spain | Maria Teresa Periñan, Pilar Gómez Garre, Pablo Mir |
| University Hospital Germans Trias i Pujol, Barcelona, Spain | Pau Pastor |
| Hospital Clínic de Barcelona, Barcelona, Spain | Ruben Fernandez-Santiago |
| Faculty of medicine university of Khartoum , Khartoum, Sudan | Sarah El-Sadig |
| Lund University, Lund, Sweden | Kajsa Brolin, Maria Swanberg |
| Karolinska Institute, Stockholm, Sweden | Per Svenningsson |

| Affiliation | Contributor |
|---|---|
| Inselspital Bern, University of Bern, Bern, Switzerland | Christiane Zweier, Gerd Tinkhauser, Paul Krack |
| National Taiwan University Hospital, Taipei City, Taiwan | Chin-Hsien Lin, Ruey-Meei Wu |
| Chang Gung Memorial Hospital, Taoyuan City, Taiwan | Hsiu-Chuan Wu |
| National Taiwan University, Taipei City, Taiwan | Pin-Jui Kung |
| Chang Gung Memorial Hospital, Taoyuan City, Taiwan | Yihru Wu |
| Avicenna Tajik State Medical University, Dushanbe, Tajikistan | Ganieva Manizha |
| National Institute Mongi Ben Hamida of Neurology, Tunis, Tunisia | Rim Amouri |
| Mongi Ben Hmida National Institute of Neurology, Tunis, Tunisia | Samia Ben Sassi |
| Koç University, Istanbul, Turkey | A. Nazlı Başak, Özgür Öztop Çakmak, Sibel Ertan |
| Şişli Etfal Training and Research Hospital, Istanbul, Turkey | Gencer Genc |
| Queen Mary University of London, London, United Kingdom | Alastair Noyce, Sumit Dey, Spencer Finch |
| University College London, London, United Kingdom | Alejandro Martínez-Carrasco, Anette Schrag, Anthony Schapira, Eleanor J. Stafford, Henry Houlden, Huw R Morris, John Hardy, Kin Ying Mok, Kin Ying Mok, Mie Rizig, Nicholas Wood, Olaitan Okunoye, Rauan Kaiyrzhanov, Rimona Weil, Simona Jasaityte, Vida Obese, Mina Ryten, Thomas Warner, Raquel Real |
| University of Plymouth, Plymouth, United Kingdom | Camille Carroll |
| Parkinson's UK, London, United Kingdom | Claire Bale |
| University of Glasgow, Glasgow, United Kingdom | Donald Grosset |
| Cardiff University, Cardiff, United Kingdom | Nigel Williams, Valentina Escott-Price |
| Royal Veterinary College University of London, London, United Kingdom | Patrick Alfryn Lewis |
| University of Bristol, Bristol, United Kingdom | Seth Love |
| Cure Parkinson's, London, United Kingdom | Simon Stott |
| St George's, University of London, London, United Kingdom | Hamin Lee |
| University of Cambridge, Cambridge, United Kingdom | Roger Barker, Caroline Williams-Gray |
| University of Oxford, Oxford, United Kingdom | Michele Hu, Laura Parkkinen |
| Northumbria Healthcare at NHS Foundation Trust, Newcastle-upon-Tyne, United Kingdom | Richard Walker |
| Imperial College London, London, United Kingdom | Steve Gentleman, Christian Lambert, Yen Tai |
| Newcastle University, Newcastle-upon-Tyne, United Kingdom | David Burn, Chris Morris |
| YLD, London, United Kingdom | Deborah Attuah |
| University of Cincinnati, Cincinnati, OH, USA | Alberto Espay, Luca Marsili |
| The Michael J. Fox Foundation for Parkinson's Research, New York, NY, USA | Alyssa O'Grady, Bernadette Siddiqi, Bradford Casey, Brian Fiske, Charisse Comart, Justin C. Solle, Kaileigh Murphy, Maggie Kuhl, Naomi Louie, Sohini Chowdhury, Todd Sherer, Ryan Pfingst, Debi Brooks, Zach Chaney, Conor Hennessey, Cassandra Barrett |

| Affiliation | Contributor |
| --- | --- |
| National Institute on Aging, Bethesda, MD, USA | Andrew B Singleton, Hampton Leonard, Kate Andersh, Laurel Screven, Kensuke Daida |
| Augusta University / University of Georgia Medical Partnership, Augusta, GA, USA | Andrew K. Sobering |
| Mid-Atlantic Permanente Medical Group, Bethesda, MD, USA | Cabell Jonas |
| Washington University, St. Louis, MO, USA | Carlos Cruchaga, Laura Ibanez |
| National Institutes of Health, Bethesda, MD, USA | Caroline B. Pantazis, Cornelis Blauwendraat, Dan Vitale, Dena Hernandez, Faraz Faghri, Hampton Leonard, Jonggeol Jeff Kim, Mary B Makarious, Mathew Koretsky, Mike A. Nalls, Sara Bandres-Ciga, Yeajin Song, Spencer Grant, Shannon Ballard, Mark Cookson, Christine Swanson-Fischer |
| Indiana University, Bloomington, IN, USA | Claire Wegel |
| Rush University, Chicago, IL, USA | Deborah Hall |
| Kaiser Permanente, Oakland, CA, USA | Ejaz Shiamim |
| Banner Sun Health Research Institute, Sun City, AZ, USA | Geidy E. Serrano |
| Data Tecnica International, Washington, WA, USA | Hirotaka Iwaki, Kristin S. Levine |
| Michigan State University, East Lansing, MI, USA | Honglei Chen |
| Cleveland Clinic, Cleveland, OH, USA | Ignacio F. Mata, Miguel Inca-Martinez, James B Leverenz |
| Kaiser Permanente, Oakland, CA, USA | Jared Williamson |
| Baylor College of Medicine, Houston, TX, USA | Joseph Jankovic, Joshua Shulman, Chad Shaw |
| Texas Children's Hospital, Houston, TX, USA | Joshua Shulman |
| Parkinson's Foundation, Princeton, NJ, USA | Kamalini Ghosh Galvelis |
| University of Miami Miller School of Medicine, Miami, FL, USA | Karen Nuytemans |
| Beth Israel Deaconess Medical Center, Boston, MA, USA | Karl Kieburtz |
| North Shore University Health System, Chicago, IL, USA | Katerina Markopoulou |
| Institute for Neurodegenerative Disorders, New Haven, CT, USA | Kenneth Marek |
| University of Pittsburgh, Pittsburgh, PA, USA | Lana M. Chahine |
| University of Alabama at Birmingham, Birmingham, AL, USA | Lauren Ruffrage, Marissa Dean, Haydeh Payami |
| University of Maryland, Baltimore, MD, USA | Lisa Shulman |
| University of Florida, Gainesville, FL, USA | Matthew Farrer, Ashley Rawls |
| Northwestern University, Chicago, IL, USA | Megan J. Puckelwartz, Steven Lubbe |
| Northwestern University, Evanston, IL, USA | Niccolò Emanuele Mencacci, Ignacio Juan Keller Sarmiento |
| Universit of Michigan, Ann Arbor, MI, USA | Roger Albin |
| Columbia University , New York, NY, USA | Roy Alcalay |
| James J. Peters Veterans Affairs Medical Center, New York, NY, USA | Ruth Walker |
| Aligning Science Across Parkinson's, Washington, WA, USA | Sonya Dumanis, Ekemini Riley |
| University of Chicago, Chicago, IL, USA | Tao Xie |

| Affiliation | Contributor |
|---|---|
| Indiana University School of Medicine, Indianapolis, IN, USA | Tatiana Foroud |
| Sun Health Research Institution, Sun City, AZ, USA | Thomas Beach |
| Aligning Science Across Parkinson's, Baltimore, MD, USA | Dana Lewis |
| Gladstone Institutes, San Francisco, CA, USA | Shreya Menon |
| Icahn School of Medicine at Mount Sinai, New York, NY, USA | Melissa Nirenberg, Rachel Saunders-Pullman |
| Banner Health, Phoenix, AZ, USA | Sidra Aslam |
| The Queen's Medical Center, Honolulu, HI, USA | Michiko Kimura Bruno |
| LSU Health Shreveport, Shreveport, LA, USA | Elizabeth Disbrow |
| University of California, Berkeley, Berkeley, CA, USA | Randy Schekman |
| NYU Grossman School of Medicine, New York City, NY, USA | Un Kang |
| VA Puget Sound Health Care System, Seattle, WA, USA | Cyrus Zabetian |
| University of California, Los Angeles, Los Angeles, CA, USA | Beate Ritz |
| Mayo Clinic, Rochester, MN, USA | Bradley Boeve, Zbigniew K. Wszolek |
| Hue University, Huế, Vietnam | Duan Nguyen, Toan Nguyen |
| University of Zambia, Lusaka, Zambia | Masharip Atadzhanov |
